# CT-Based Deep Foundation Model for Predicting Immune Checkpoint Inhibitor-Induced Pneumonitis Risk in Lung Cancer

**DOI:** 10.64898/2026.04.21.26351428

**Authors:** Amgad Muneer, Eman Showkatian, Yuliya Kitsel, Maliazurina B. Saad, Sheeba J Sujit, Felipe Soto, Girish S. Shroff, Saadia A. Faiz, Mohammad I. Ghanbar, Sherif M. Ismail, Natalie I. Vokes, Tina Cascone, Xiuning Le, Jianjun Zhang, Lauren A. Byers, David Jaffray, Joe Y. Chang, Zhongxing Liao, Aung Naing, Don L. Gibbons, Ara A. Vaporciyan, John V. Heymach, Karthik S. Suresh, Mehmet Altan, Ajay Sheshadri, Jia Wu

## Abstract

**Background:** Immune checkpoint inhibitors (ICIs) have revolutionized cancer therapy but can cause serious immune-related adverse events (irAEs), with pneumonitis (ICI-P) being among the most severe. Early identification of high-risk patients before ICI initiation is critical for close monitoring, timely intervention, and optimizing outcomes.

**Purpose:** To develop and validate a deep learning foundation model to predict ICI-P from baseline CT scans in patients with lung cancer.

**Methods:** We designed the Checkpoint-Inhibitor Pneumonitis Hazard EstimatoR (CIPHER), a deep learning-powered foundation model combining contrastive learning with a transformer-based masked autoencoder to predict ICI-P from baseline CT scans in lung cancer patients. Using self-supervised learning, CIPHER was pretrained on 590,284 CT slices from 2,500 non-small cell lung cancer (NSCLC) patients, to learn representations of heterogeneous lung parenchyma. Following pretraining, CIPHER was adapted to the internal MDA NSCLC immunotherapy cohort of 347 patients, of whom 33 developed adjudicated ICI-P. Fine-tuning was performed using 254 non-ICI-P patients only, and a held-out internal validation set of 93 patients, including 33 ICI-P cases and 60 non-ICI-P controls, was reserved for evaluation. CIPHER was benchmarked against clinical, radiomics, and ensemble comparator models and externally validated in an independent Johns Hopkins NSCLC cohort of 116 patients, including 20 ICI-P cases and 96 non-ICI-P controls.

**Results:** In our internal immunotherapy cohort, CIPHER consistently distinguished patients at elevated risk of ICI-P from those without the event, with AUCs ranging from 0.77 to 0.85. In head-to-head benchmarking, CIPHER achieved an AUC of 0.83, outperforming the clinical, radiomics and ensemble models. In the external validation cohort, CIPHER maintained high performance (AUC=0.83; balanced accuracy=81.7%), exceeding the radiomics model (DeLong p=0.0318) and demonstrating superior specificity without sacrificing sensitivity. By contrast, the radiomics model, despite high sensitivity (85.0%), showed markedly lower specificity (45.8%). Confusion matrix analyses confirmed CIPHER’s robust classification, correctly identifying 80 of 96 non–ICI-P cases and 16 of 20 ICI-P cases.

**Conclusions:** We developed and externally validated CIPHER, a CT-based imaging biomarker for pretreatment ICI-P risk stratification in NSCLC. CIPHER shows promise as a noninvasive tool for ICI-P risk assessment but warrants prospective validation before clinical translation.

**Highlights:**

- **The first chest CT AI foundation model for immune toxicity** – We introduce CIPHER (Checkpoint-Inhibitor Pneumonitis Hazard EstimatoR), a transformer-based masked autoencoder trained through self-supervised contrastive learning on 590,284 CT slices from 4,242 CT scans from NSCLC patients. This large-scale pretraining enables CIPHER to learn intrinsic lung parenchymal representations linked to immune toxicity risk.
- **Early risk prediction prior to therapy initiation** – CIPHER predicts the likelihood of ICI-induced pneumonitis directly from *baseline* CT scans, offering the first non-invasive foundation model for early risk assessment before ICI.
- **Robust validation and benchmarking** – We fine-tuned and evaluated CIPHER across independent internal and external NSCLC immunotherapy cohorts, achieving AUCs of 0.77–0.85 internal and 0.83 external testing, surpassing comparator models in both performance and generalizability.
- **Interpretability and translational potential** – We demonstrate how model-derived attention maps align with clinically relevant pulmonary patterns, enhancing interpretability.
- **Translational potential** – CIPHER’s performance and scalability underscore its potential as a decision-support tool to guide treatment planning, pre-emptive monitoring, and toxicity mitigation in immunotherapy practice.

## INTRODUCTION

Immune checkpoint inhibitors (ICIs) have dramatically transformed the landscape of cancer treatment, particularly in non-small cell lung cancer (NSCLC), which accounts for 85% of all lung cancers ^1–6^. By blocking inhibitory checkpoints like PD-1 and CTLA-4, ICIs activate immune cells to target and destroy tumor cells, improving survival in many treatment settings ^7^. However, these benefits come with the risk of immune-related adverse events (irAEs) ^8^ ^9^, which arise when unrestrained T-cell activation disrupts immune homeostasis and peripheral self-tolerance in healthy organs ^10^ ^11^.

Among these irAEs, ICI-induced pneumonitis (ICI-P) is particularly concerning due to its potential for severe, life-threatening lung inflammation^12–14^. ICI-P occurs in approximately 10% of NSCLC patients receiving ICIs, and it is an independent predictor of increased morbidity and non–cancer-related mortality in this population ^15^ ^16^. Despite its clinical significance, reliable tools for identifying patients at high risk for developing ICI-P are currently lacking ^16–20^. As a result, identifying patients at high risk for ICI-P is challenging, thus complicating the development of prospective trials to reduce ICI-P risk in these patients.

Emerging evidence suggests that the risk for ICI-P is not uniformly distributed among patients. Pre-existing conditions, such as interstitial lung disease (ILD) or interstitial lung abnormalities (ILAs), have been associated with a higher risk of developing ICI-P ^21^ ^22^. However, these require interpretation from expert thoracic radiologists, and are often reported qualitatively. To address this challenge, we developed a novel deep learning foundation model, CIPHER (Checkpoint-Inhibitor Pneumonitis Hazard EstimatoR), to predict the risk of developing ICI-P using baseline CT scans. The primary goal of this study is to identify patients at high risk for ICI-P, which may pave the way for future trials focused on screening, monitoring, and preventive or pre-emptive interventions.

## METHODS

### Study Definitions

The primary outcome was ICI-related pneumonitis, defined by compatible respiratory symptoms (e.g., cough, dyspnea) and concordant CT abnormalities. CT patterns were categorized by the dominant abnormality as nonspecific interstitial pneumonitis (NSIP), organizing pneumonia (OP), hypersensitivity pneumonitis (HP), diffuse alveolar damage (DAD), or indeterminate, and severity was graded using the latest Common Terminology Criteria for Adverse Events (CTCAE) v5.0^23–25^. The primary outcome of interest was grade ≥ 2 (CTCAEv5). The same clinical-radiologic adjudication framework was used in the MDA and Johns Hopkins cohorts, and adjudicators were blinded to CIPHER scores, reconstruction-error maps, and model classifications. Grade 1 pneumonitis was not included in the event endpoint. A multidisciplinary adjudication panel of pulmonologists, thoracic radiologists, thoracic oncologists, and infectious disease specialists reviewed each case using clinical history, imaging, laboratory and microbiological data, and treatment response to immunosuppressive therapies to determine whether findings were attributable to pneumonitis versus alternative etiologies (e.g., infection or progression).

### Study Cohorts

This retrospective study was approved by our institutional review board and was conducted in compliance with the Health Insurance Portability and Accountability Act (HIPAA) and the ethical standards of the 1964 Declaration of Helsinki and its later amendments. Patient consent was waived for this study.

#### The University of Texas MD Anderson Cancer Center (MDA) Lung Cancer Foundation Model Cohort

We used a cohort of 2,500 NSCLC patients with 4,242 thoracic CT scans to pretrain the CIPHER foundation model to learn lung anatomical structures. This cohort was derived by sampling about 12.5% of patients from our institutional GEMINI database, which includes over 20,000 patients with lung cancer treated at MDA. The goal of this pre-training phase was to enable the model to learn diverse lung cancer anatomy patterns before being applied to predict ICI-P risk. The dataset reflected real-world clinical imaging diversity, with technical heterogeneity in acquisition parameters and scanner models. Nearly half of the CT scans were acquired at outside hospitals using a diverse range of scanners and imaging protocols, as reported in our previous study^26^, thereby simulating the heterogeneity seen in multi-institutional datasets. Most scans had an in-plane matrix size of 512 × 512 and a median slice thickness of 2.5 mm. This variability was systematically addressed through standardized preprocessing and incorporated into model training to enhance robustness.

#### MDA Immunotherapy Cohort

We analyzed pre-treatment CT scans from 347 NSCLC patients treated with ICIs between 2013 and 2021, either as monotherapy or in combination with chemotherapy from the GEMINI cohort^27^. Among these, 33 patients developed ICI-P, as adjudicated by a multi-disciplinary review team. Clinical and imaging data, including demographics, treatment history, and radiologic findings, were retrospectively collected from the electronic health records. Patients were excluded if baseline chest CT scans were unavailable or of poor diagnostic quality. Further details about the cohort have been described elsewhere^27^. CT scans were obtained using various scanners and protocols; most were performed with GE Medical Systems scanners (74%), followed by Siemens (23%) and Toshiba (3%), and axial plane pixel spacing had a median of 0.8 mm (0.4 - 1.4 mm), and slice thickness had a median of 2.5 mm (0.2 - 5 mm).

#### Johns Hopkins Immunotherapy Cohort (JHU)

The independent external validation cohort consisted of 116 NSCLC patients who received ICIs at Johns Hopkins Hospital. Inclusion criteria were consistent with those of the MDA cohort. 20 patients developed ICI-P and 96 did not; ICI-P was adjudicated by a multidisciplinary clinical team. CT scans were acquired using various scanners – Siemens (71.9%), Toshiba (22.2%), and GE (5.9%). Acquisition parameters also varied, with a median slice thickness of 2.5 mm (range: 0.2–10 mm) and a median axial pixel spacing of 0.75 mm (range: 0.5–5 mm). Patient flow and cohort construction are presented in **supplemental Figure S1**.

### Preprocessing

All CT scans underwent standardized preprocessing prior to model development. Images were first cropped to the foreground region and resampled to a standardized voxel spacing of 1.5 × 1.5 × 2.0 mm. Intensity values were clipped using a lung-specific window (−330 to 150 HU) to suppress non-informative extremes and enhance parenchymal contrast. During training, random spatial sampling was applied to improve robustness and mitigate overfitting. Automated lung segmentation was performed using the LungMask framework^28^, a deep learning–based approach employing pretrained U-Net models for both whole-lung and lobar segmentation. Outputs from these models were combined to generate a robust binary lung mask, which was subsequently used to constrain downstream analysis to pulmonary regions. Tumor regions, atelectasis, obstructive changes, and lymphangitic spread within the lung mask were not manually excluded. For model input, volumetric data were partitioned into 3D blocks of size 96 × 96 × 96 voxels, enabling efficient learning of spatially contextual imaging features.

### Overall Study Design

We hypothesized that a deep learning-based approach could objectively detect and quantify such abnormalities from pre-treatment CT scans to improve ICI-P risk prediction. To test this hypothesis, we developed CIPHER, a foundation model trained using self-supervised learning. This approach was chosen to enable the model to learn comprehensive representations of lung parenchyma from a large cohort of lung cancer patients. CIPHER was trained to reconstruct lung anatomy; abnormal regions such as ILD/ILA were expected to produce high reconstruction errors. These reconstruction errors were then used as a quantitative imaging biomarker for predicting risk of ICI-P. The study contained four critical steps (**Fig 1**) including foundation model development, task-specific adaptation, comparative evaluation, and external validation.

**Figure 1:**
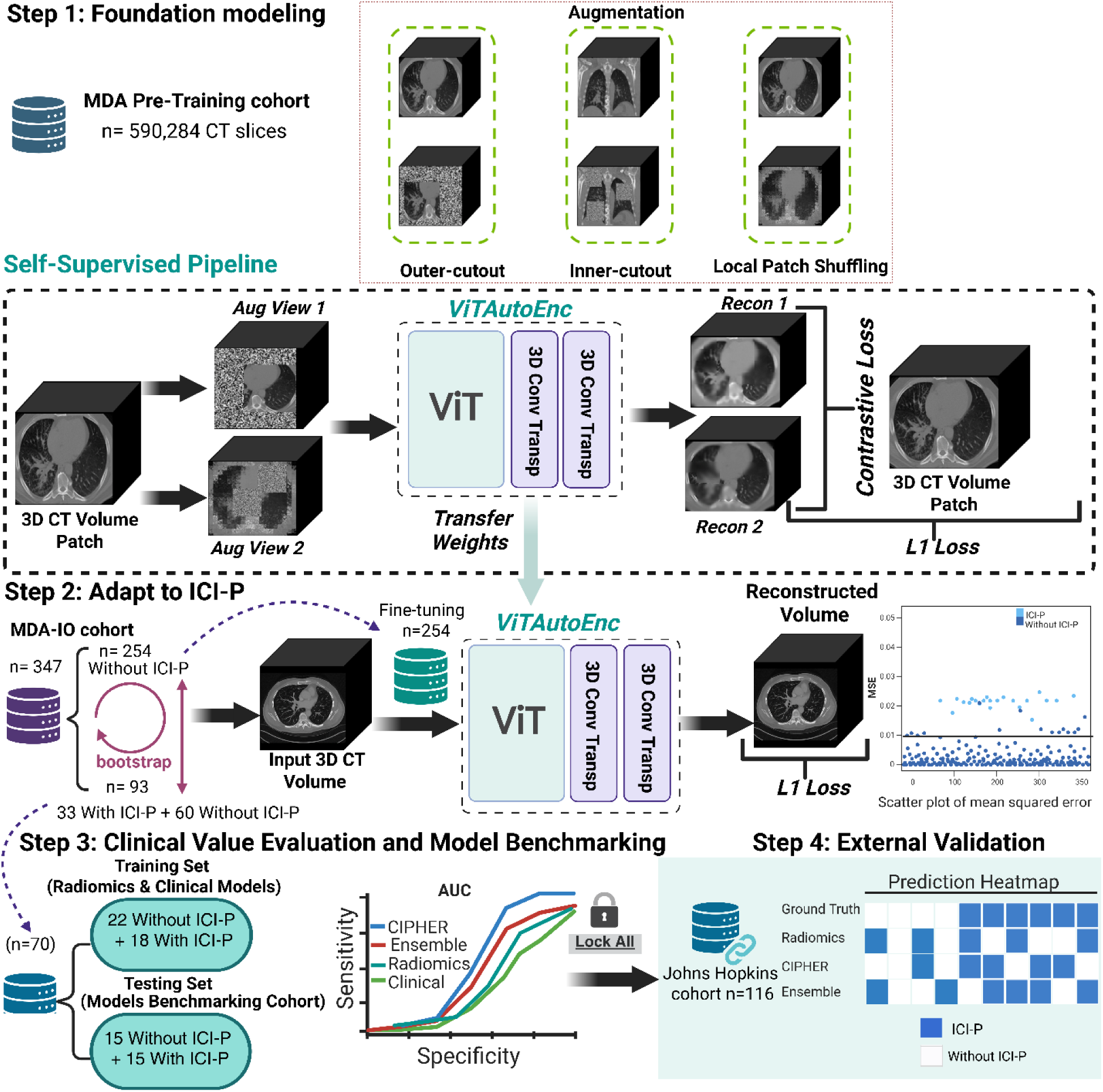
Overview of the CIPHER foundation model for predicting ICI-P from baseline CT scans in NSCLC. The model is pretrained on 590,284 CT slices using a self-supervised 3D Vision Transformer autoencoder (Step 1). It is then adapted to the ICI-P task by fine-tuning on 254 non–ICI-P patients and evaluated on a hold-out cohort of 93 patients (33 ICI-P, 60 non–ICI-P), where the reconstruction error was summarized as a patient-level ICI-P risk score (Step 2). Model performance is benchmarked against clinical, radiomics, and ensemble comparator models in an internal MDA benchmark cohort (Step 3), and the final locked models are externally validated in an independent JHU cohort (n=116) to assess generalizability (Step 4).

### Step 1: Construction of CT Foundation Model

To address the challenge of accurately predicting a rare event like ICI-P from a limited number of annotated scans, we developed CIPHER, a deep learning foundation model (**Fig 1**). We adopted a self-supervised learning (SSL) framework to leverage the large, unlabeled CT dataset, enabling the model to learn robust and generalizable representations of thoracic anatomy prior to fine-tuning for the specific prediction task^29^. This two-stage approach is particularly well-suited here, where unlabeled imaging data are abundant, but expert annotation is resource-intensive^30^.

The backbone architecture of CIPHER is a transformer-based masked autoencoder^31^. A transformer architecture was selected over conventional Convolutional Neural Networks (CNNs) because of its superior capacity to model long-range spatial dependencies and capture global contextual information—features critical for detecting diffuse, subtle parenchymal abnormalities that may signal early ICI-P risk^32^.

The model was pre-trained on 590,284 unique 2D axial slices from 4,242 3D CT scans. Following the masked autoencoder principle, random image patches were masked, and the encoder learned a compact latent representation from which a lightweight decoder reconstructed the original image^31^. This mask and reconstruction task forced the model to learn a deep contextual and anatomical understanding without explicit labels. To enhance both representational fidelity and discriminative power, a dual-objective loss function was used during pre-training (details in the **Supplemental Methods**). Optimization targeted both a contrastive loss and L1 reconstruction loss, the former enhancing the discriminative capability of learned representations, and the latter preserving high-fidelity image reconstruction. Additional implementation details for CIPHER are provided to support reproducibility (**Supplementary Table S1, Fig S2**).

### Step 2: Fine-tuning Foundation Model for ICI-P Risk Prediction

Following pre-training, CIPHER was fine-tuned on a subset of the MDA immunotherapy cohort and subsequently locked for testing on hold-out patients. Specifically, 254 cases without ICI-P were used for fine-tuning, and the remaining 93 patients (33 with ICI-P, 60 without) were reserved for CIPHER internal evaluation. CIPHER was developed as a self-supervised reconstruction-derived imaging risk model rather than a conventional supervised binary classifier. Therefore, ICI-P and non-ICI-P class labels were not used jointly during CIPHER pretraining or fine-tuning, and no positive/negative class balancing was performed for CIPHER. This phase enabled CIPHER to adapt its learned thoracic anatomy representations for the specific task of ICI-P risk prediction. During fine-tuning, model parameters were refined to reconstruct the original CT scans in detecting subtle imaging patterns using only L1 loss. For risk stratification, the reconstruction error between each patient’s original CT scan and the corresponding autoencoder-reconstructed image was used as the imaging risk score. This error reflects regions in which the model was unable to fully reconstruct the underlying anatomical information. Reconstruction error was calculated within the segmented lung volume and summarized at the patient level to generate the CIPHER imaging risk score for subsequent developing adjudicated ICI-P. A single mean squared error (MSE)-based classification threshold was selected using the internal development data and locked before evaluation. The locked threshold was then applied unchanged to both the internal held-out cohort and the external validation cohort to classify patients into low- and high-risk groups. Further methodological details are provided in the **Supplemental Materials**. To assess reproducibility, a resampling strategy was employed, repeating this process five times with non-overlapping subsets of non–ICI-P patients used for fine-tuning and testing in each iteration.

### Step 3: Benchmarking Foundation Model

The internal analyses served distinct purposes. Five subsampling runs were used to assess CIPHER performance on a hold-out cohort of 93 patients, including 33 ICI-P and 60 non-ICI-P patients. Separately, for head-to-head benchmarking against supervised comparator models, we defined a balanced internal MDA benchmark cohort of 30 patients, including 15 ICI-P and 15 non-ICI-P patients. This benchmark cohort was used to compare CIPHER with supervised clinical, radiomics, and ensemble models under the same test conditions, while the remaining MDA cases were used for supervised comparator training. The radiomics model was developed using a set of 107 standard CT features extracted with PyRadiomics (v2.2.0). The radiomics model was trained using AutoGluon^33^, an automated machine learning framework that selects and optimizes state-of-the-art models. The under-sampling strategy^34^ was used only for supervised comparator models training to mitigate class imbalance by reducing the number of majority-class cases to match the minority-class sample size, where positive and negative labels were used during training, and it was not applied to CIPHER. The ensemble model was constructed by combining the predicted probabilities from the clinical, radiomics, and CIPHER models using a weighted-average approach.

To prevent data leakage, all downstream model development and evaluation were performed at the patient level. The self-supervised pretraining cohort was separate from the MDA immunotherapy cohort used for CIPHER fine-tuning and internal evaluation; no patient, scan, or slice from the internal held-out evaluation set was included in pretraining. The external JHU cohort was entirely unseen during pretraining, fine-tuning, threshold selection, and model evaluation.

### Step 4: External Validation

To assess generalizability, the CIPHER model and comparator models were locked and applied without retraining or fine-tuning to an independent cohort of 116 NSCLC patients from the Johns Hopkins immunotherapy cohort. The same preprocessing and analysis pipeline used for the MDA cohort was employed to ensure a fair, direct comparison of model performance across institutions.

### Statistical Analysis

Model performance was assessed using balanced accuracy, specificity, sensitivity, F1 score, the area under the receiver operating characteristic curve (AUC), positive predictive value (PPV), and negative predictive value (NPV). The optimal threshold for classifying patients into high- and low-risk groups for subsequent adjudicated ICI-P was determined in the internal development data using Youden’s J statistic and then locked before evaluation. This threshold was applied unchanged to the internal held-out cohort and the external JHU validation cohort. Calibration was assessed using Brier scores and calibration plots, and clinical utility was evaluated using decision-curve analysis. The statistical significance of the multivariate model (i.e., whether its AUC exceeded that of a random classifier, AUC = 0.5) was evaluated using the Mann–Whitney U test. The DeLong test was used to calculate 95% confidence intervals for AUCs and to compute p-values for pairwise comparisons of ROC curves. Associations between CIPHER outputs and CTCAE pneumonitis grade were assessed using Spearman correlation, Kruskal-Wallis tests, and Mann-Whitney U tests. Time to ICI-P was evaluated using Cox proportional hazards models, Kaplan-Meier curves, and log-rank tests. All statistical tests were two-sided, with p < 0.05 considered statistically significant.

## RESULTS

### Characteristics of Immunotherapy Cohorts

A total of 347 patients treated between 2013 and 2021 were included in this study. Detailed demographic and clinical characteristics of the cohort are presented in **Table 1**. The MDA model development cohort had a median age of 65 years and included 181 (52%) males and 166 (48%) females. The majority of patients were White/Caucasian (n=269, 77%). The cohort had a significant history of tobacco usage; 70% were former smokers and 10% were current smokers. Among patients with a smoking history, a heavy burden of exposure was common, with 44.9% having smoked for more than 40 pack-years. Pre-existing lung conditions were prevalent, with 202 patients (58%) having a prior diagnosis of chronic obstructive pulmonary disease (COPD) and 6 (2%) having clinically evident interstitial lung disease (ILD) at baseline. At the time of ICI initiation, the most frequently reported symptoms were cough (n=94, 27%) and shortness of breath (n=86, 25%).

**Table 1:**
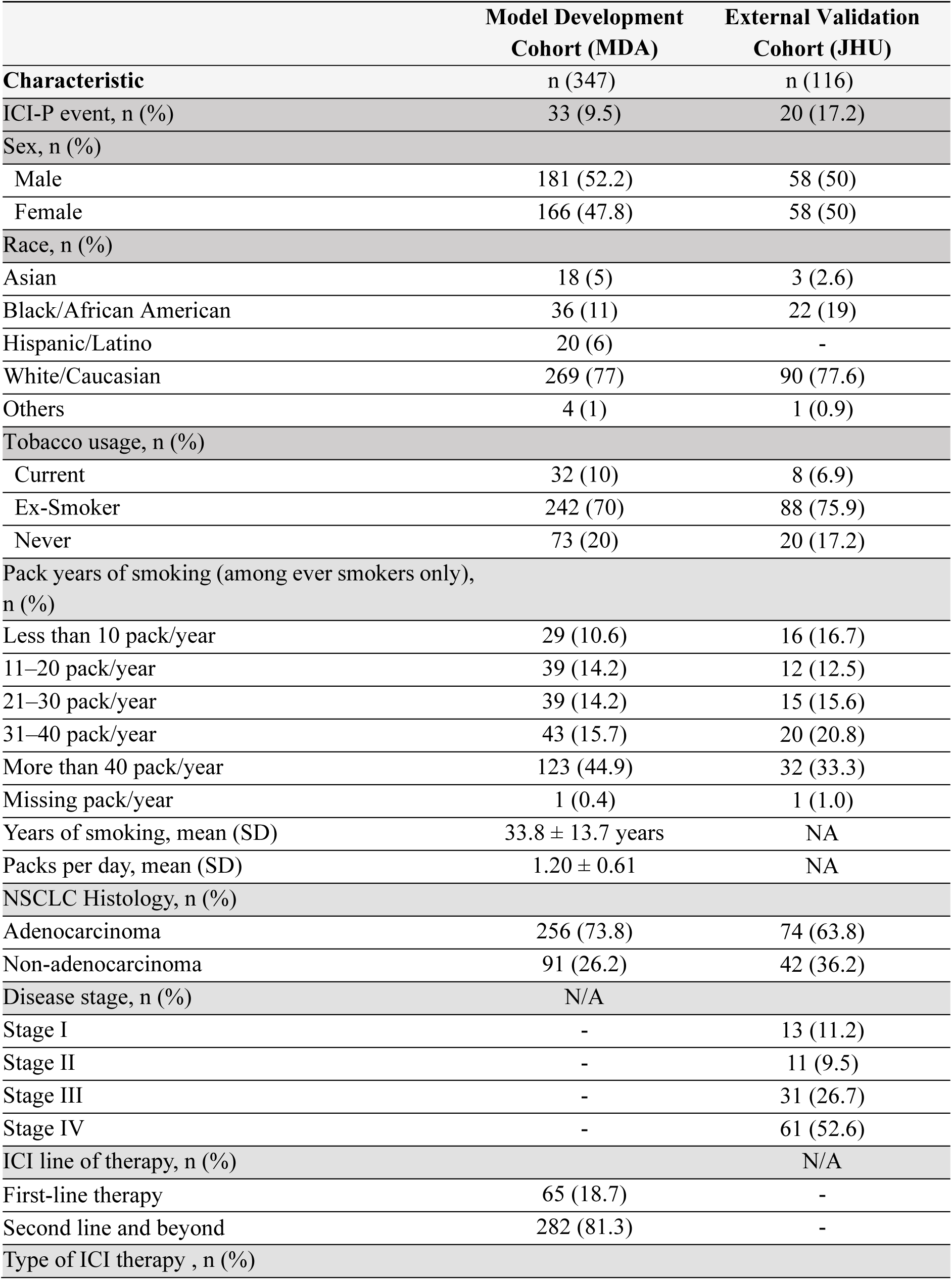
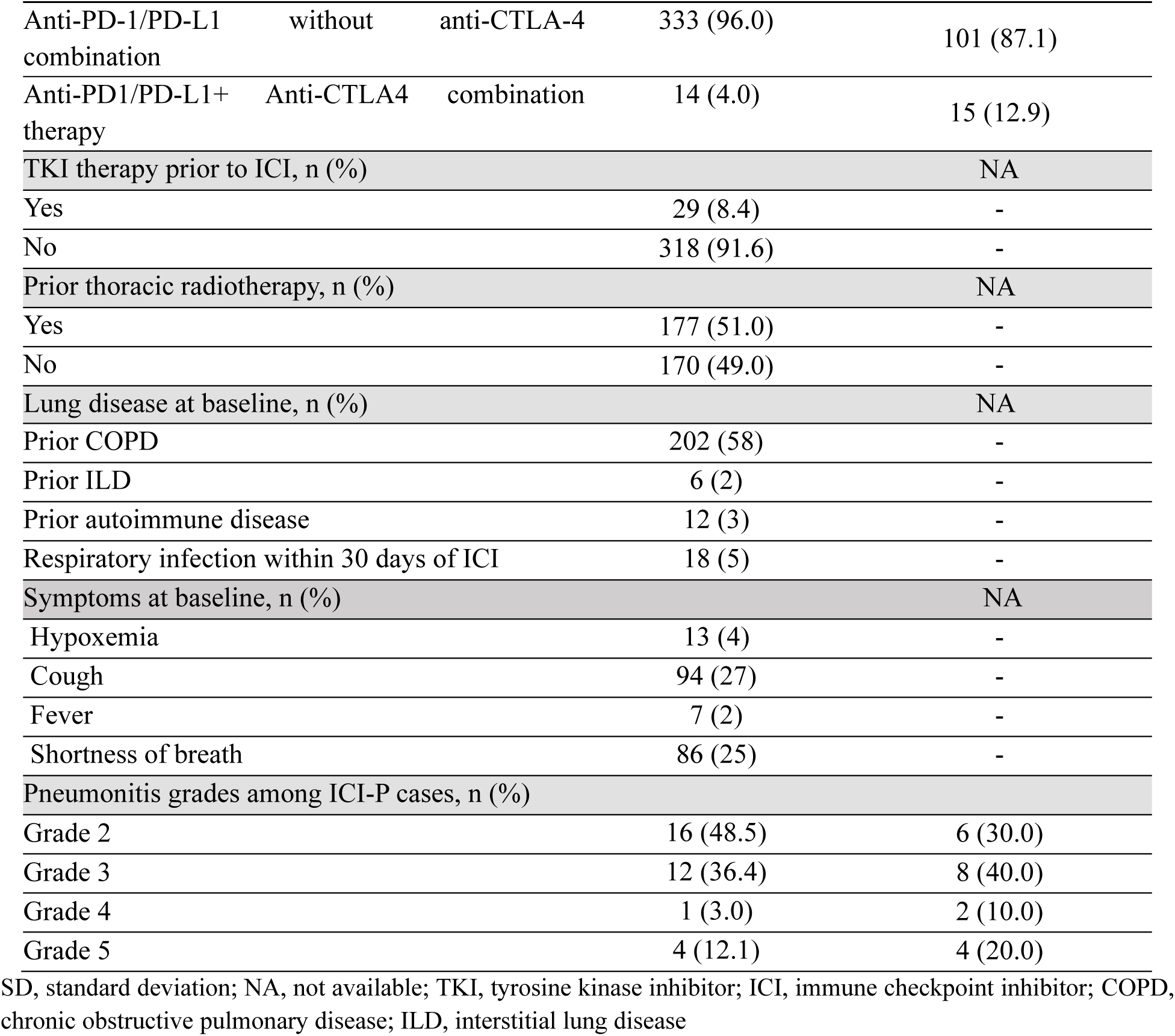
Demographics and clinical characteristics of the study immunotherapy cohorts.

In the external validation cohort from Johns Hopkins (n=116), the mean age at ICI initiation was 65 years (SD ± 9.7), with an equal distribution of males (n=58, 50%) and females (n=58, 50%). The majority of patients were White/Caucasian (n=90, 77.6%). Tobacco exposure was also common; 75.9% were ex-smokers, 6.9% were current smokers, and 17.2% were never smokers. The most common reported pack-year category was more than 40 pack-years (n=32, 33.3%), followed by 31–40 pack-years (n=20, 20.8%) and less than 10 pack-years (n=16, 16.7%); pack-year data were missing for one patient (1.0%). Symptoms at baseline, detailed chemotherapy-combination and antiangiogenic-combination variables, prior TKI exposure, and detailed radiation variables were not uniformly available in the JHU cohort.

Additional tumor and treatment characteristics are summarized in Table 1. NSCLC histology was available in both cohorts. In the MDA cohort, 256 of 347 patients, 73.8%, had adenocarcinoma and 91, 26.2%, had non-adenocarcinoma histology. In the JHU cohort, 74 of 116 patients, 63.8%, had adenocarcinoma and 42, 36.2%, had non-adenocarcinoma histology. While the MDA cohort only included patients with metastatic disease, the JHU cohort included 13 stage I cases, 11.2%; 11 stage II cases, 9.5%; 31 stage III cases, 26.7%; and 61 stage IV cases, 52.6%. In the MDA cohort, ICI was administered as first-line therapy in 65 patients, 18.7%, and as second-line or later therapy in 282 patients, 81.3%. Anti-PD-1/PD-L1 therapy without anti-CTLA-4 combination was used in 333 MDA patients, 96.0%, and 101 JHU patients, 87.1%; anti-PD-1/PD-L1 plus anti-CTLA-4 combination therapy was used in 14 MDA patients, 4.0%, and 15 JHU patients, 12.9%. Prior TKI exposure before ICI was available only in the MDA cohort and was present in 29 patients, 8.4%. Disease stage in the MDA cohort, line of therapy and prior TKI exposure in the JHU cohort, and driver mutation status and PD-L1 expression in both cohorts were not available in the curated datasets.

Because prior thoracic radiotherapy may confound ICI-P risk assessment and baseline CT interpretation, we performed radiation-specific analyses in the MDA cohort. Prior thoracic radiotherapy was common, occurring in 177 of 347 patients, 51.0%. It was present in 19 of 33 ICI-P cases, 57.6%, and 158 of 314 non-ICI-P patients, 50.3%. Prior thoracic radiotherapy was not significantly associated with ICI-P in the full MDA cohort, OR = 1.34, 95% CI 0.65-2.77, Fisher exact p = 0.468. In the 93-patient internal held-out cohort, CIPHER probability remained significantly associated with ICI-P after adjustment for prior thoracic radiotherapy, adjusted OR per 0.1 increase = 3.00, 95% CI 1.94-4.65, p < 0.001. Prior thoracic radiotherapy was not independently associated with ICI-P after accounting for CIPHER probability, adjusted OR = 1.38, 95% CI 0.47-4.05, p = 0.563. No statistically significant CIPHER-by-radiotherapy interaction was observed, p = 0.113.

We then further characterized the 19 ICI-P cases with prior thoracic radiotherapy. Among irradiated ICI-P cases with available dates, the median interval between thoracic radiotherapy and ICI initiation was 10 days before ICI initiation, range 0-1096 days. Available treatment-field information confirmed lung parenchymal involvement. Among 17 irradiated ICI-P cases with sufficient field-distribution information, no cases were confined entirely within the prior radiation field; 6 of 17 cases, 35.3%, occurred outside the prior radiation field, and 11 of 17 cases, 64.7%, involved both prior radiation-field and out-of-field lung regions. Complete dosimetry maps and spatially registered treatment fields were not uniformly available, precluding quantitative voxel-level colocalization of CIPHER attention or reconstruction-error maps with irradiated lung regions.

Endpoint adjudication was performed for 72 suspected pneumonitis cases in the MDA cohort. After review, 39 cases were excluded because an alternative diagnosis was favored, leaving 33 adjudicated grade ≥2 ICI-P cases. Among these 33 cases, CTCAE grade distribution was 16 grade 2 cases, 48.5%; 12 grade 3 cases, 36.4%; 1 grade 4 case, 3.0%; and 4 grade 5 cases, 12.1%. Radiologic pattern information was available for all 33 adjudicated MDA ICI-P cases and included OP in 14 cases, 42.4%; NSIP in 12 cases, 36.4%; DAD/AIP-ARDS in 6 cases, 18.2%; and HP in 1 case, 3.0%. No cases were classified as bronchiolitis or indeterminate. In the JHU external validation cohort, CTCAE grade distribution included 6 grade 2, 8 grade 3, 2 grade 4, and 4 grade 5 ICI-P cases.

### CIPHER Model Performance on MDA Immunotherapy Cohort

CIPHER can effectively distinguish between patients without ICI-P and those at elevated risk. In the violin plot (**Fig 2a**), patients without ICI-P (blue dots) and those at risk (orange crosses) are clearly separated by the threshold (black line); most cases with low reconstruction errors were ICI-P–negative, while those with high reconstruction errors were at higher predicted risk, with Mann–Whitney U test yielded U = 15.0, with a highly significant result (p < 0.0001). Receiver-operating characteristic curve (ROC) analysis (**Fig 2b**) yielded area under the curve (AUC) values ranging from 0.77 to 0.85 across five independent subsampling runs (p<0.001), indicating robust and consistent performance. Confusion matrices for each run (**Fig 2c**) provide additional details on classification outcomes. The associated performance metrics, including balanced accuracy, sensitivity, specificity, AUC, and F1 score are summarized for each subsampling run in **supplemental Table S2**.

**Figure 2:**
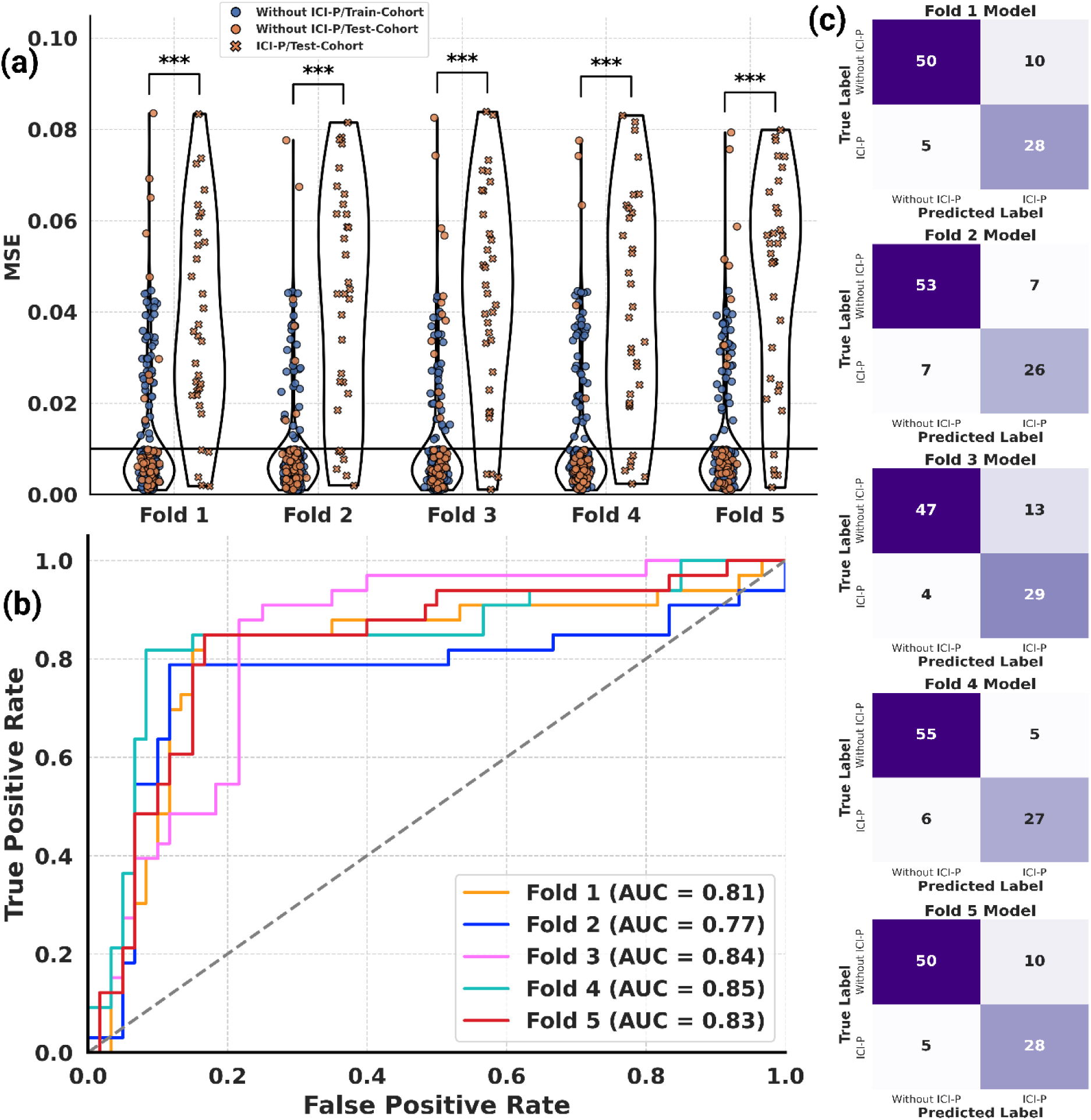
Performance evaluation of the CIPHER foundation model on MDA cohort: (a) Violin plot for ICI-P and non-ICI-P cohorts across five subsampling runs; (b) ROC curves showing AUC scores for each fold; and (c) Confusion matrices for five subsampling runs indicating prediction accuracy.

### Benchmarking CIPHER Against Comparator Models on the MDA Immunotherapy Cohort

We evaluated clinical, radiomics, CIPHER, and ensemble models on a reserved MDA internal benchmark cohort of 15 ICI-P and 15 non–ICI-P cases for fair head-to-head comparison. ROC curves (**Fig 3a**) confirmed the performance ranking: CIPHER achieved the highest AUC among the evaluated models (AUC =0.83, 95% CI 0.663-0.990), compared with the clinical model (AUC = 0.58, 95% CI 0.367-0.793), radiomics model (AUC = 0.77, 95% CI 0.592-0.955), and ensemble model (AUC = 0.78, 95% CI 0.605-0.951). In pairwise DeLong comparisons, CIPHER had a higher AUC than the clinical, radiomics, and ensemble models, although these differences did not reach statistical significance: CIPHER versus clinical model, p = 0.0759; CIPHER versus radiomics model, p = 0.6257; and CIPHER versus ensemble model, p = 0.4962 (**Supplementary Table S3**).

**Figure 3:**
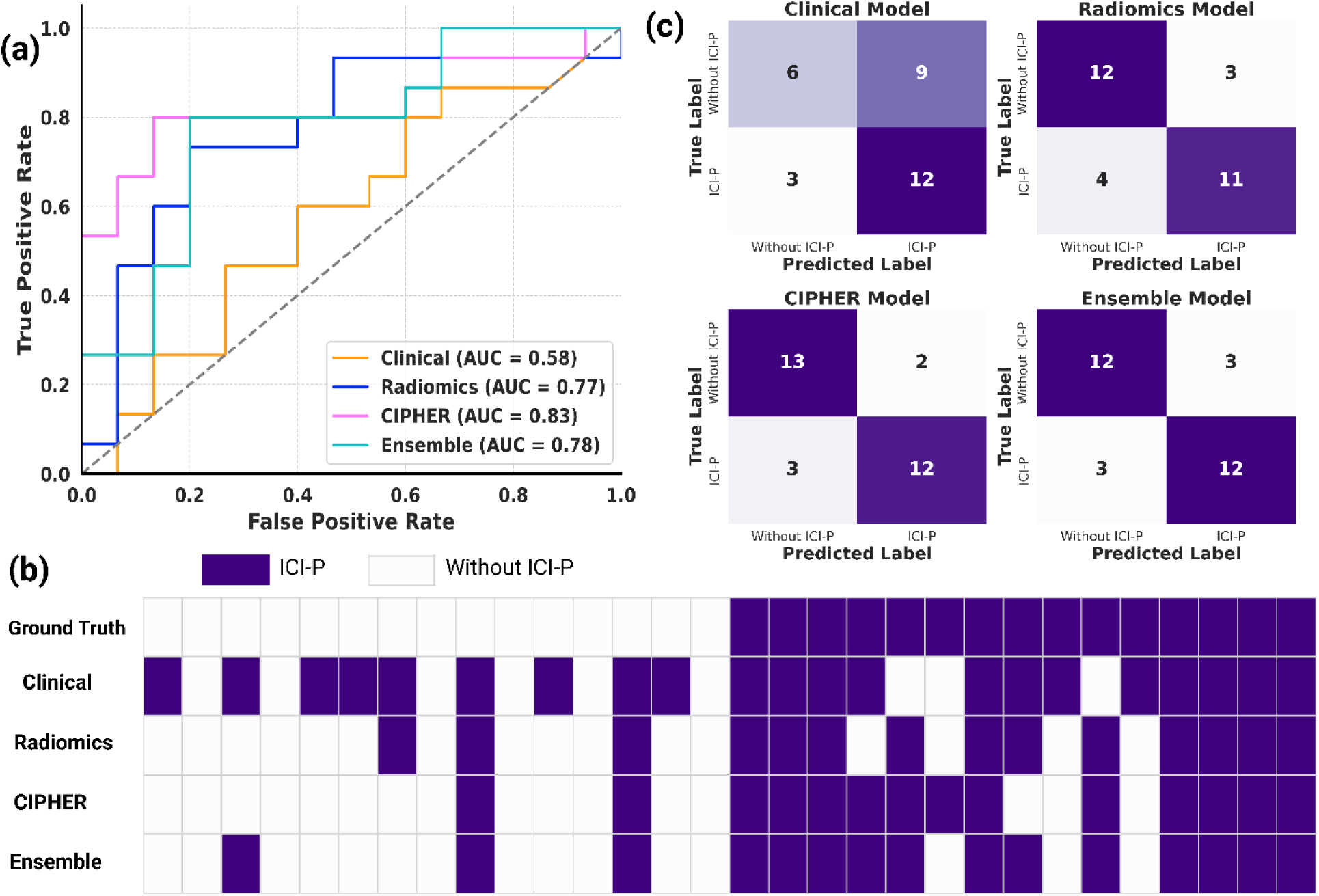
Performance comparison of clinical, radiomics, CIPHER, and ensemble models for ICI-P prediction on MDA internal benchmark cohort: (a) ROC curves benchmarking; (b) Binary heatmap of ground truth versus predicted labels showcasing the closer alignment of CIPHER model predictions with actual outcomes; and (c) Confusion matrices illustrate the prediction results for each model.

Predicted vs. actual label comparisons (**Fig 3b**) showed close agreement for the CIPHER model with the ground truth labels. Confusion matrices (**Fig 3c**) revealed that the clinical model correctly classified 6 of 15 non–ICI-P cases and 12 of 15 ICI-P cases, the radiomics model correctly classified 12 of 15 non–ICI-P cases and 11 of 15 ICI-P cases, CIPHER correctly classified 13 of 15 non–ICI-P cases and 12 of 15 ICI-P cases, and the ensemble model correctly classified 12 of 15 non–ICI-P cases and 12 of 15 ICI-P cases. Performance metrics summarized in **Table 2** showed that CIPHER achieved the highest balanced accuracy (83.3%) and specificity (86.7%) while maintaining sensitivity of 80.0% and F1 score of 82.8%. CIPHER also achieved the highest PPV, 85.7%, with an NPV of 81.3%, compared with PPV/NPV of 57.1%/66.7% for the clinical model, 78.6%/75.0% for the radiomics model, and 80.0%/80.0% for the ensemble model. The ensemble model achieved balanced sensitivity and specificity of 80.0%, but did not improve AUC or balanced accuracy beyond CIPHER alone.

**Table 2:** Performance of predictive models in MDA internal benchmark cohort and external validation cohorts.

| Model | B Accuracy | Specificity | Sensitivity | F1 Score | AUC (95% CI) | PPV | NPV |
| --- | --- | --- | --- | --- | --- | --- | --- |
| Internal Benchmark Cohort (MDA) |  |  |  |  |  |  |  |
| Clinical | 60.0 | 40.0 | 80.0 | 66.7 | 58.0<br>(0.367-0.793) | 57.1 | 66.7 |
| Radiomics | 76.7 | 80.0 | 73.3 | 75.9 | 77.3<br>(0.592-0.955) | 78.6 | 75.0 |
| CIPHER | 83.3 | 86.7 | 80.0 | 82.8 | 82.7<br>(0.663-0.990) | 85.7 | 81.3 |
| Ensemble | 80.0 | 80.0 | 80.0 | 80.0 | 77.8<br>(0.605-0.951) | 80.0 | 80.0 |
| External Validation Cohort (JHU) |  |  |  |  |  |  |  |
| Clinical | 64.1 | 78.1 | 50.0 | - | 58.7<br>(0.437-0.738) | 32.3 | 88.2 |
| Radiomics | 65.4 | 45.8 | 85.0 | - | 66.3<br>(0.551-0.776) | 24.6 | 93.6 |
| CIPHER | 81.7 | 83.3 | 80.0 | - | 82.6<br>(0.713-0.938) | 50.0 | 95.2 |
| Ensemble | 76.2 | 62.5 | 90.0 | - | 79.6<br>(0.695-0.897) | 33.3 | 96.8 |

Additionally, calibration and clinical-utility analyses further supported the performance of CIPHER in the MDA internal benchmark cohort. CIPHER had the lowest Brier score among the evaluated models, 0.187, compared with 0.248 for the clinical model, 0.200 for the radiomics model, and 0.203 for the ensemble model (**Supplementary Fig S3, Table S4)**. Decision-curve analysis using model-predicted probabilities showed a favorable net-benefit profile for CIPHER. CIPHER provided higher net benefit than the clinical model across most evaluated thresholds and showed a favorable profile compared with the radiomics and ensemble models over much of the clinically relevant range (**Supplementary Fig S3)**. Prevalence-adjusted PPV and NPV analyses were also performed across clinically plausible ICI-P prevalence levels to account for expected variation in predictive values across practice settings (**Supplementary Table S5)**.

### CIPHER Maintains Highest Performance in External Validation on JHU Immunotherapy Cohort

We evaluated clinical, radiomics, CIPHER, and ensemble models on an independent Johns Hopkins cohort (n=116, **Table 1**) to assess external validation performance. The violin plot for CIPHER (**Fig 4a**) demonstrated clear separation between ICI-P and non–ICI-P cases (p < 0.001), with higher prediction errors in true positives, reflecting confident risk identification. ROC analysis (**Fig 4b**) confirmed CIPHER’s robust performance, achieving an AUC of 0.83, 95% CI 0.713-0.938, compared with the clinical model (AUC=0.59, 95% CI 0.437-0.738), radiomics model (AUC=0.66, 95% CI 0.551-0.776), and ensemble model (AUC=0.80, 95% CI 0.695-0.897). Pairwise DeLong testing showed that CIPHER significantly outperformed the clinical model, p = 0.0287, and the radiomics model, p = 0.0318, while CIPHER and the ensemble model had comparable AUCs, p = 0.5996 (**Supplementary Table S3**).

**Figure 4:**
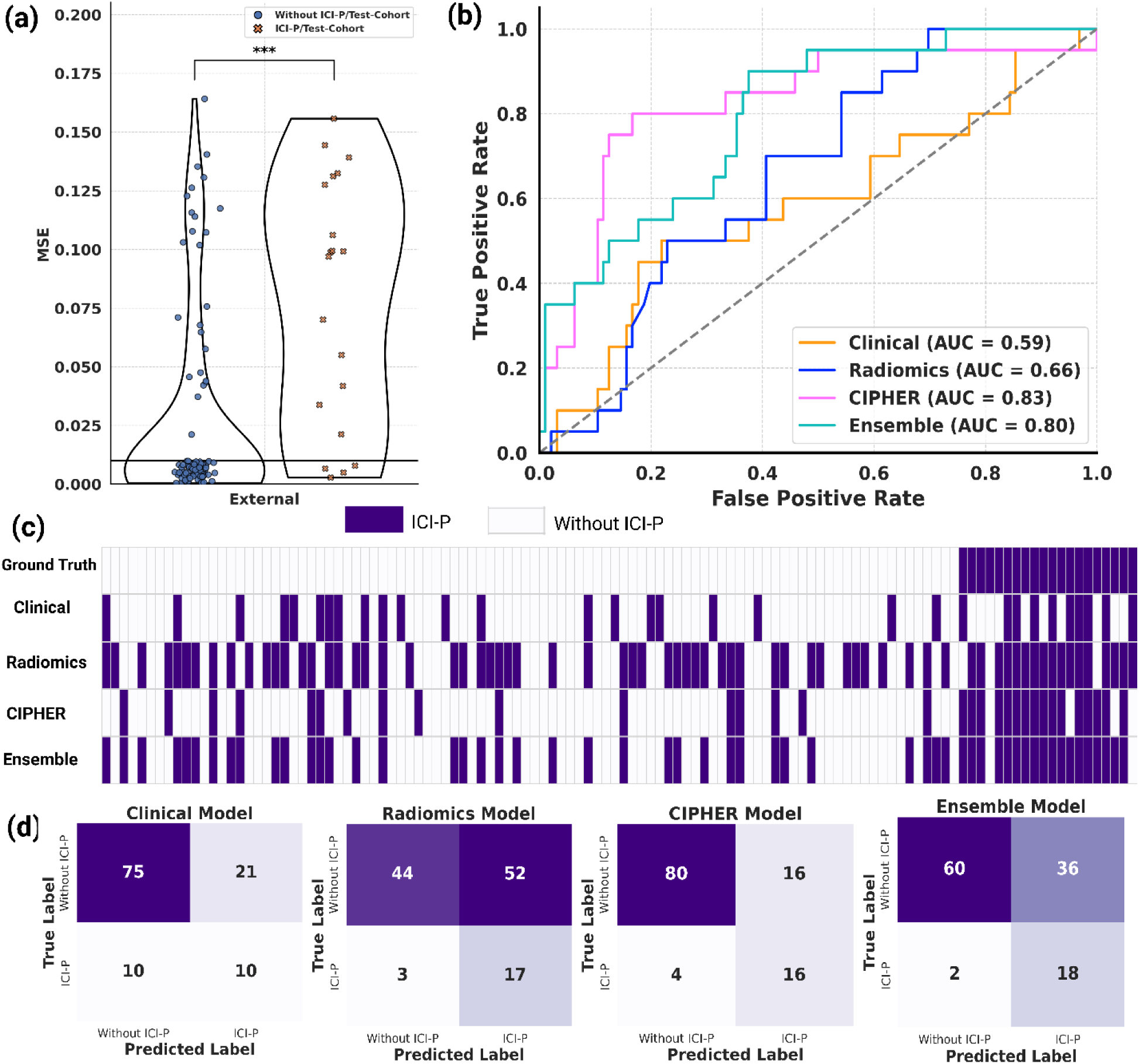
Performance evaluation of the CIPHER foundation model on external cohort: (a) Violin plot for ICI-P and non-ICI-P cohorts. The statistical significance is indicated as *** = p < 0.001. (b) ROC curves showing AUC scores for each model, (c) Binary heatmap showing ground truth vs. predictions of different models, and (d) Confusion matrices.

Case-level prediction plots (**Fig 4c**) further illustrate CIPHER’s predictions closely aligned with ground truth, outperforming clinical and radiomics models and showing fewer false-positive classifications than the ensemble model. Confusion matrices (**Fig 4d**) further supported the robust performance of CIPHER, which correctly classified 16 out of 20 ICI-P cases and 80 out of 96 negatives. As summarized in **Table 2**, CIPHER achieved the highest balanced accuracy (81.7%), exceeding the clinical model (64.1%), radiomics model (65.4%), and ensemble model (76.2%). CIPHER also maintained balanced sensitivity (80.0%) and specificity (83.3%). CIPHER achieved the highest PPV (50.0%) and a high NPV (95.2%), compared with PPV/NPV of 32.3%/88.2% for the clinical model, 24.6%/93.6% for the radiomics model, and 33.3%/96.8% for the ensemble model. In contrast, the radiomics model, despite high sensitivity (85.0%), showed poor specificity (45.8%) with a high false-positive rate. The ensemble model achieved the highest sensitivity (90.0%) but lower specificity (62.5%) and balanced accuracy (76.2%) than CIPHER, indicating increased detection of ICI-P at the cost of additional false-positive classifications.

Supplementary calibration and clinical-utility analyses showed that CIPHER had the lowest Brier score in the JHU external validation cohort, 0.175, compared with 0.215 for the clinical model, 0.237 for the radiomics model, and 0.194 for the ensemble model. Decision-curve analysis showed that CIPHER provided higher net benefit than the clinical and radiomics models across the evaluated threshold range and exceeded the ensemble model over most thresholds. Prevalence-adjusted PPV and NPV analyses across clinically plausible ICI-P prevalence levels are provided in Supplementary Table S5. Full results are provided in Supplementary Figure S3, Tables S4 and S5.

### Robustness, Sensitivity, and Ablation Analyses

Sensitivity analyses were performed in the MDA internal held-out cohort to evaluate whether CIPHER performance was driven by known baseline ILD or overt baseline pulmonary abnormalities. After excluding patients with known baseline ILD, CIPHER maintained similar performance, with an AUC of 0.839 compared with 0.845 in the primary internal held-out cohort. Similar results were observed after excluding patients with known ILD or restrictive/combined pulmonary impairment, with an AUC of 0.839. In a broader exploratory analysis excluding patients with known ILD or documented baseline pulmonary abnormality, CIPHER showed lower but persistent discrimination, with an AUC of 0.762. These findings suggest that CIPHER performance was not solely driven by known baseline ILD, while also supporting baseline lung vulnerability as part of the imaging risk signal. Full performance metrics are provided in **Supplementary Table S6**.

We also evaluated whether CIPHER performance was driven by recent respiratory infection or baseline respiratory symptoms in the MDA internal held-out cohort. Performance remained stable after excluding patients with recent respiratory infection within 30 days of ICI initiation, AUC 0.851, after excluding patients with baseline fever, AUC 0.841, and after excluding patients with any baseline respiratory symptom, AUC 0.853. In the most stringent analysis excluding patients with recent infection, fever, or any baseline respiratory symptom, CIPHER maintained discrimination, with an AUC of 0.838, balanced accuracy of 0.868, sensitivity of 0.813, and specificity of 0.923. These findings suggest that CIPHER performance was not explained solely by recent infection or baseline respiratory symptoms. Further metrics are provided in **Supplementary Table S7**.

Additionally, we performed a sensitivity analysis to evaluate whether CT contrast status influenced CIPHER performance in the JHU external validation cohort. Among 116 patients, 98, 84.5%, had contrast-enhanced CT scans and 18, 15.5%, had non-contrast CT scans. CIPHER maintained strong performance in the contrast-enhanced subgroup, with an AUC of 0.826, sensitivity of 0.789, specificity of 0.722, PPV of 0.405, and NPV of 0.934. In the non-contrast subgroup, CIPHER achieved an AUC of 0.941; however, this subgroup included only 18 patients and 1 ICI-P event and was therefore interpreted cautiously. No significant interaction was observed between CIPHER score and contrast status for ICI-P prediction, p = 0.456, suggesting no clear evidence that CT contrast status affected CIPHER performance in the external cohort. Full model-specific results are provided in **Supplementary Table S8**.

### CIPHER Stratifies Time to ICI-P in the Internal MDA Test Cohort

We next performed a secondary time-to-event analysis to evaluate whether CIPHER was associated with earlier ICI-P onset after ICI initiation. Among the 33 MDA patients who developed ICI-P, the median interval between baseline CT and ICI initiation was 19 days, IQR 7-37 days, and the median time from ICI initiation to pneumonitis was 187 days, IQR 83-328 days, range 6-867 days. In the internal test cohort, higher CIPHER probability was significantly associated with shorter time to ICI-P. Each 0.1 increase in CIPHER probability was associated with increased ICI-P hazard, HR 2.31, 95% CI 1.63-3.28, p <0.001. This association remained significant using conservative censoring at the earliest available treatment stop, progression, death, or last follow-up date, HR 1.93, 95% CI 1.37-2.73, p <0.001. Kaplan-Meier analysis showed earlier ICI-P occurrence in the CIPHER high-risk group than in the low-risk group, log-rank p < 0.001 (**Fig 5**). External time-to-event analysis was not performed because ICI initiation dates and censoring dates for non-ICI-P controls were not uniformly available in the deidentified Johns Hopkins cohort.

**Figure 5:**
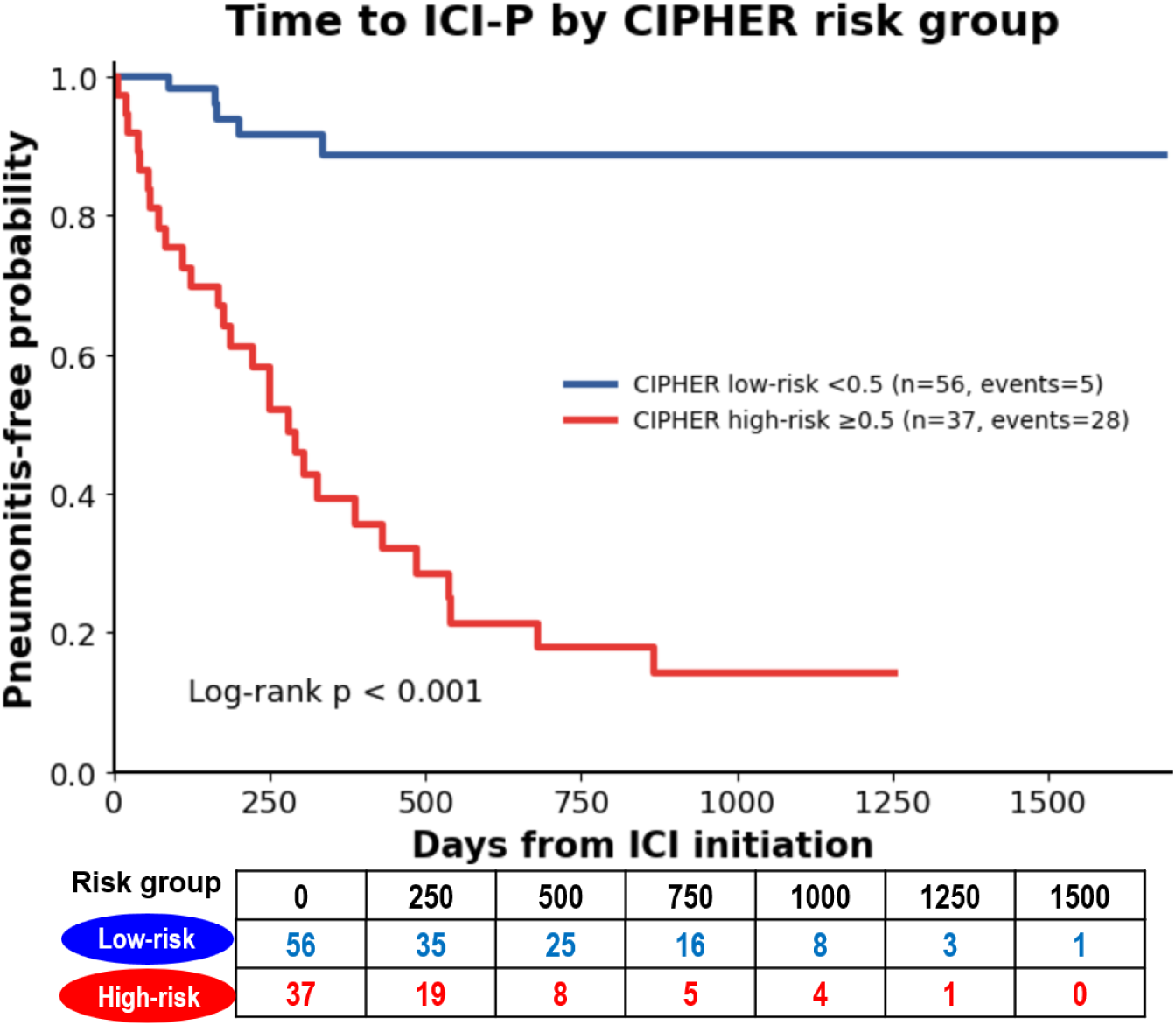
Time to ICI-P by CIPHER risk group in the internal test cohort. Kaplan-Meier curves show pneumonitis-free probability from ICI initiation, stratified by the CIPHER probability threshold. The CIPHER high-risk group had earlier ICI-P occurrence than the low-risk group, log-rank p < 0.001. The x-axis includes both ICI-P event times and censoring times; therefore, the curve extends beyond the maximum observed ICI-P event time because some non-ICI-P patients had longer follow-up. The number-at-risk table is shown below the plot.

### Exploratory Association of CIPHER Outputs with CTCAE Pneumonitis Grade

We performed exploratory analyses to evaluate whether CIPHER outputs were associated with CTCAE pneumonitis grade. These analyses were intended to assess whether CIPHER identified ICI-P risk across clinically relevant severity levels, not to develop or validate a CTCAE grade classifier. In the MDA internal held-out cohort, the grade distribution included 16 grade 2, 12 grade 3, 1 grade 4, and 4 grade 5 ICI-P cases. Using the locked CIPHER threshold, CIPHER detected 13 of 16 grade 2 cases, 81.2%; 10 of 12 grade 3 cases, 83.3%; 1 of 1 grade 4 case, 100.0%; and 4 of 4 grade 5 cases, 100.0%. CIPHER probability showed a modest association with CTCAE grade in the internal cohort, Spearman ρ = 0.359, p = 0.040, with Kruskal-Wallis p = 0.030, whereas CIPHER MSE was not significantly associated with grade, Spearman ρ = 0.042, p = 0.816, with Kruskal-Wallis p = 0.973 (**Supplementary Table S9, Fig S4**).

In the JHU external validation cohort, the grade distribution included 6 grade 2, 8 grade 3, 2 grade 4, and 4 grade 5 ICI-P cases. CIPHER detected 4 of 6 grade 2 cases, 66.7%; 8 of 8 grade 3 cases, 100.0%; 2 of 2 grade 4 cases, 100.0%; and 2 of 4 grade 5 cases, 50.0%. Neither CIPHER predicted probability nor CIPHER MSE was significantly associated with CTCAE grade in the external cohort. For predicted probability, Spearman ρ = 0.367, p = 0.111, with Kruskal-Wallis p = 0.255; for MSE, Spearman ρ = −0.285, p = 0.223, with Kruskal-Wallis p = 0.463. These analyses were exploratory and were not intended to evaluate multiclass CTCAE grade prediction (**Supplementary Table S9, Fig S4**).

### CIPHER Remained a Strong Independent Predictor After Adjusting for Clinical Context on Two-Center Data

The CIPHER scores were strong predictors of ICI-P in univariate analysis and after adjustment, while other clinical variables were not (**Fig 6**). In MDA cohort, each one-standard-deviation increase in CIPHER corresponded to approximately a three-fold increase in the risk of toxicity. Baseline fever was inversely associated with risk of ICI-P in unadjusted, but not adjusted models. Other demographic or clinical variables, including age, sex, histology, and smoking status, were not significantly associated with ICI-P after adjustment (**Fig 6a**). Similar trends were observed in external JHU validation cohort. In univariate analyses, the CIPHER probability score showed the strongest association with ICI-P, and remained significant after multivariable adjustment, indicating that CIPHER provided independent predictive information beyond clinical and demographic factors such as age, race, and smoking status (**Fig 6b**).

**Figure 6:**
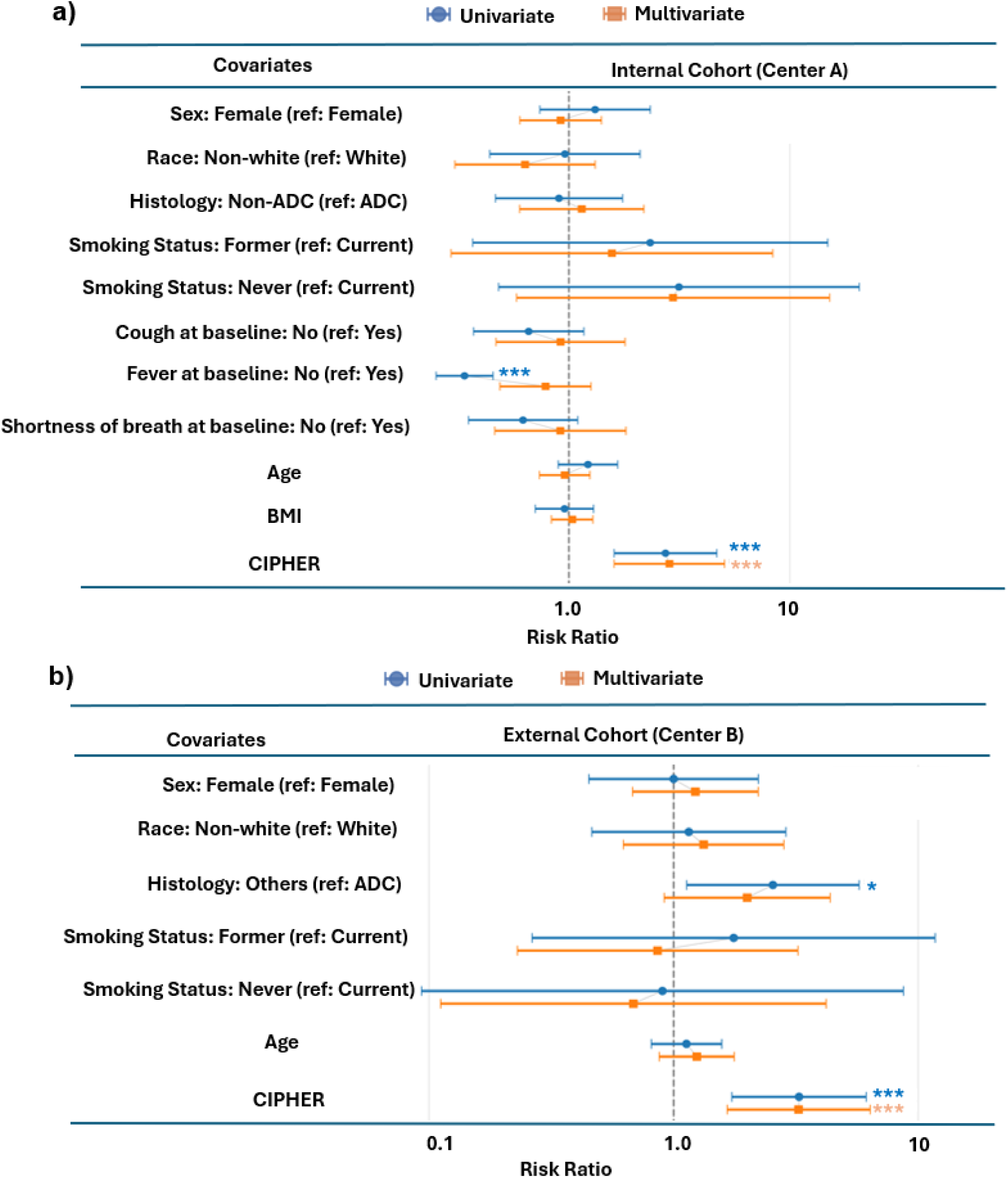
Forest plots summarizing univariate and multivariate risk ratio analyses for the CIPHER foundation model. (a) Internal MDA testing cohort (n = 93) and (b) External JHU validation cohort (n = 116). Each plot displays risk ratios on a log scale with 95% confidence intervals across demographic, clinical, and model-derived features. Statistical significance is indicated as *p < 0.05, **p < 0.01, and ***p < 0.001

### Interpretation of CIPHER Model

The findings illustrated in **Fig 7** highlight the clinical interpretability and predictive potential of the CIPHER model in identifying patients at risk for ICI-P using baseline CT scans. The model localized high-risk pulmonary regions prior to the onset of pneumonitis, as shown by the attention maps in **Fig 7 (middle)**. Across representative cases, the attention maps consistently emphasized regions with interstitial abnormalities, particularly fibrotic-appearing patterns such as subpleural reticulation and ground-glass opacities (white arrows), which may reflect pre-existing and potentially undiagnosed interstitial lung disease. Notably, the model also assigns high attention to non-fibrotic findings in some instances, including atelectatic/consolidative opacities (green arrows) and vascular structures (blue arrow), suggesting potential confounding signals or correlated imaging markers. Collectively, these visualizations support that CIPHER leverages clinically meaningful pulmonary features from the baseline CT.

**Figure 7:**
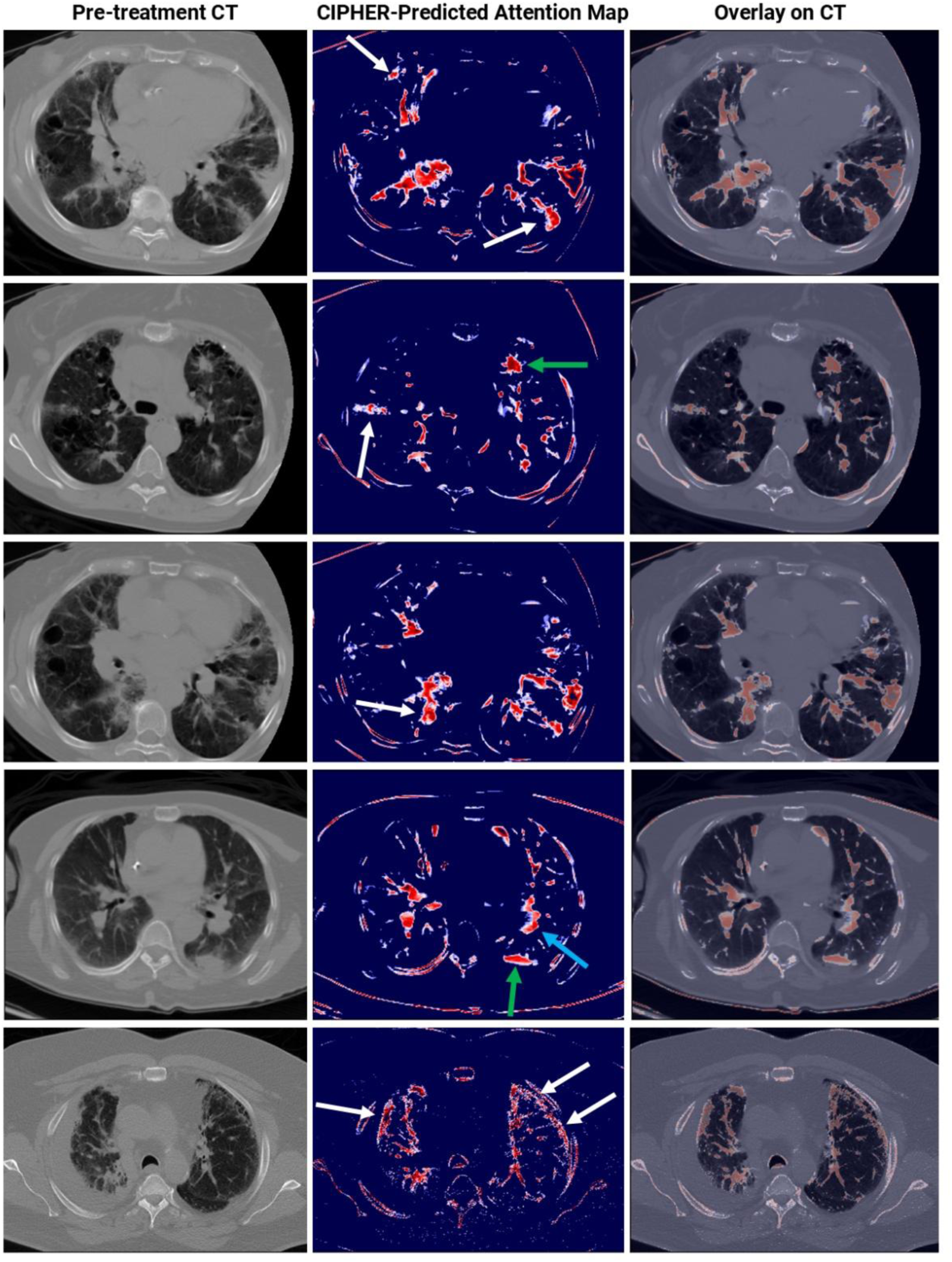
Visualization of CIPHER model attention in interstitial lung disease. Representative examples of pre-treatment CT scans (left), CIPHER-predicted attention maps (middle), and overlays on CT (right). Red areas denote high model attention that in most instances corresponds to fibrotic interstitial abnormalities (white arrows) including subpleural reticulation and ground-glass opacities. In some instances, however, high model attention areas correspond to non-fibrotic abnormalities including atelectatic/consolidative opacities (green arrows) and vessels (blue arrow).

To further evaluate interpretability, we performed a quantitative clinical/radiologic correlation and error analysis in the internal held-out cohort. CIPHER correctly classified 28 of 33 ICI-P cases and 51 of 60 non-ICI-P controls, with 9 false-positive and 5 false-negative cases. We compared CIPHER MSE scores across baseline clinical/radiologic variables using the Mann-Whitney U test. Higher CIPHER MSE scores were associated with baseline hypoxemia (median 0.0634 vs 0.0069, p=0.030) and baseline cough (median 0.0091 vs 0.0062, p=0.046), with a trend toward higher scores in patients with COPD (median 0.0091 vs 0.0059, p=0.069). We also performed a descriptive error analysis of misclassified cases. False-positive cases frequently had potential confounding lung-risk features, including prior thoracic radiation (6/9, 66.7%), COPD (5/9, 55.6%), baseline cough (4/9, 44.4%), and baseline shortness of breath (3/9, 33.3%). False-negative cases were also commonly associated with prior thoracic radiation (4/5, 80.0%). These findings suggest that CIPHER risk scores may capture clinically relevant baseline pulmonary vulnerability, while also highlighting potential confounding by pre-existing lung disease and treatment-related lung injury.

From a translational perspective, CIPHER should be interpreted as a pretreatment imaging risk-stratification aid, not as a standalone diagnostic or treatment-decision tool. In a potential clinical workflow, a routine baseline chest CT obtained before ICI initiation would undergo standardized preprocessing and lung segmentation, followed by CIPHER-based generation of a patient-level risk score, risk category, and visual explanation maps. These outputs could support multidisciplinary review by oncology, thoracic radiology, and pulmonary medicine when clinically indicated. For patients classified as high risk, CIPHER could support baseline pulmonary assessment, review for ILD/ILA or radiation-related lung injury, optimization of comorbid lung disease, patient counseling about early pneumonitis symptoms, closer symptom surveillance, earlier follow-up imaging, and a lower threshold for pulmonary consultation. Low-risk patients would continue standard guideline-based monitoring.

## DISCUSSION

In this study, we demonstrate that a novel deep learning foundation model, CIPHER, can accurately predict patients with NSCLC at high risk for ICI-P using pre-treatment chest CT scans. Compared with alternative clinical and radiomics models, CIPHER achieved superior predictive performance. Although further validation in larger, multi-center datasets is essential, our pilot findings suggest that CIPHER could facilitate personalized monitoring strategies and preventive interventions for patients most vulnerable to ICI-P.

ICI-P is a potentially life-threatening toxicity of ICI therapies, which are standard of care for the frontline treatment of NSCLC without actionable genetic alterations and other malignancies.^13^ Though risk factors for ICI-P have been published, these often vary from study to study and depend on qualitative interpretation of radiographic imaging and clinical characteristics.^14^ ^35^ ^36^ CIPHER offers an unbiased tool for ICI-P risk stratification and enables the selection of a high-risk cohort that could be enrolled into studies that mitigate ICI-P risk by selectively modulating inflammatory pathways, reducing the risk for irAEs without compromising anti-tumor immunity. Conversely, patients predicted to be at low risk could be monitored less aggressively, reducing both patient burden and healthcare costs.^37^ Though vigilant monitoring is essential for ICI-treated patients, the ability to identify those at the highest risk before therapy starts could enable a paradigm shift toward proactive management.^38^

To our knowledge, this is the first study to build a CT-based foundation model to investigate irAEs. Our study adds onto prior radiomics and radiogenomics studies,^18^ ^39–53^ to unlock latent prognostic information embedded in routine chest CT scans, as a window into lung health. Unlike existing radiomics studies, which depend on supervised learning on labor-intensive annotated datasets, we used a fundamentally different foundation model design. Consistent with emerging evidence, foundation models can capture higher-order imaging phenotypes beyond human-defined descriptors, offering a leap in precision over conventional radiomics studies^18^ ^39–47^ ^54^.

Specifically, we employed self-supervised learning on large unlabeled datasets to reconstruct lung parenchyma from a diverse lung cancer population. Because ILD or ILAs—established risk factors for ICI-P—are relatively rare and exhibit heterogeneous manifestations, the model may reconstruct these abnormalities less accurately. As a result, the degree of reconstruction error reflects the extent of pre-existing parenchymal abnormality, which serves as a predictor for elevated ICI-P risk. Further work is needed to understand the clinical significance of abnormal lung parenchyma and how they increase the risk for ICI-P.

This framework also enables seamless integration of multimodal data to further improve the tool. For example, blood-based biomarkers, like Krebs-von den Lungen-6 (KL-6)^55^, a biomarker of ILD that also had a modest ability to predict ICI-P in NSCLC^56^, could be incorporated into the CIPHER framework to improve prediction. In addition, adding positron emission tomography (PET)-derived metabolic data may further enhance prediction, as metabolic changes can precede structural abnormalities on CT and help distinguish inflammatory from fibrotic processes.^57^ While these could improve the current version of the tool, our data show that CIPHER performs well even when using only information from pre-treatment chest CT.

Our current work has limitations that guide future model development. We focused on NSCLC, which has the highest risk and incidence rate of ICI-P among solid organ tumors. However, the approach can be extended to other cancers. Although the CIPHER model was validated externally on the JHU dataset, false negatives remain a critical concern. In addition, attention maps and reconstruction-error patterns should be interpreted as model-derived imaging correlates of ICI-P risk, not as causal mechanisms. Radiologist-derived baseline CT scoring was not uniformly available, limiting assessment of CIPHER’s incremental value beyond expert visual evaluation. Thus, future studies with larger, multi-institutional cohorts are needed to improve performance and assess the clinical utility of imaging-based predictors. On the technical side, advanced self-supervised learning architectures may boost model performance by learning richer latent imaging representations.

In summary, we have developed and externally validated CIPHER, a foundation model capable of predicting future risk of developing ICI-P from pre-treatment CT scans. With future prospective validation, CIPHER can be incorporated into routine pre-ICI assessments, enabling clinicians to stratify patients into tailored monitoring pathways and identify optimal candidates for preventive intervention trials.

## Data Availability

We confirm that the code and data supporting this study's findings are openly available, as reported in this article, and its supplementary materials. The code repository for this work is supplied here: [GitHub: https://github.com/WuLabMDA/CIPHER], while data used in this study will be accessible to the research community upon reasonable request from the corresponding author.

## DECLARATIONS

### Ethics approval and consent to participate

The requirement was waived by IRB 2021-0123 for MDA cohort and the IRB 00186276 for JHU cohort due to the retrospective nature of the study.

### Consent for publication

Not applicable.

### Availability of data and materials

The CIPHER code repository is available at https://github.com/WuLabMDA/CIPHER and includes inference code, preprocessing scripts, example configuration files, instructions for preparing input CT scans, an example case for demonstration, and example outputs illustrating the expected CIPHER risk-score and visualization formats. The trained CIPHER model weights are not included in the public repository. Deidentified data and trained model weights may be made available from the corresponding author upon reasonable request, subject to institutional approval, and data-use agreements.

### Authors’ contributions

A.M., E.S., A.S., and J.W. conceptualized the clinical problem and overall study design. A.M., E.S., M.B.S., Y.K., S.J.S., F.S., G.S.S., S.A.F., M.I.G., and S.M.I. acquired and processed the multicenter data. A.M. and E.S. designed and analyzed the CIPHER model and prepared the initial manuscript draft. E.S., A.S., J.W., and A.M. revised the model development and validation framework across multisite cohorts. A.M., E.S., A.S., and J.W. developed and trained the models and performed the statistical analyses for the final version. A.M., E.S., A.S., and J.W. performed quality control of the data and algorithms. A.S., M.A., and J.W. supervised the project. M.I.G., N.I.V., T.C., X.L., J.Z., L.A.B., D.J., J.Y.C., Z.L., A.N., D.L.G., A.A.V., J.V.H., K.S.S., M.A., A.S., and J.W. contributed to manuscript review and editing. All authors had access to the data presented in the manuscript and approved the final version for publication.

