## Supplemental Material for "CT-Based Deep Foundation Model for Predicting Immune Checkpoint Inhibitor-Induced Pneumonitis Risk in Lung Cancer"

### Supplemental Methods

We optimized two loss functions during this stage to ensure the model learned meaningful and discriminative features. Contrastive loss encouraged augmented views from the same 3D CT block to produce similar feature representations while separating representations from different blocks within the same minibatch. L1 loss minimized the difference between reconstructed and original CT blocks, promoting high-fidelity image reconstruction (*Supplementary A*). The CIPHER training and validation loss metrics exhibit rapid convergence, with a significant reduction over 1000 epochs, indicating effective model learning (***Suppl Figure S2a***). The model's contrastive loss and reconstruction loss metrics further validate the quality of learned representations and reconstruction accuracy, respectively (***Suppl Figure S2b and S2c***). The weights learned during this SSL training were then transferred to the next phase of CIPHER model development. The following are the two-loss and augmentation methods employed in our foundation model development.

*A. Foundation Model Loss*

Our study utilized two key loss functions: Contrastive Loss and L1 Loss. These loss functions are essential in training the CIPHER model, ensuring robust representation learning and accurate reconstruction of 3D CT volume blocks.

***Contrastive Loss***

The contrastive loss function[1] is adapted to ensure that the representations of augmented views derived from the same original 3D CT volume block are similar, while those derived from different blocks within the same minibatch are dissimilar. Given a minibatch of $N$ 3D CT volume blocks, each original block $x_i$ undergoes augmentation, resulting in two views $x_i^{(1)}$ and $x_i^{(2)}$. The similarity between the representations of these views $z_i^{(1)}$ and $z_i^{(2)}$ is measured using cosine similarity:

$$\mathrm{sim}(\mathrm{u},\mathrm{v}) = \frac{\mathrm{u}^{\top}\mathrm{v}}{\| \mathrm{u} \| \| \mathrm{v} \|}$$

The contrastive loss for a positive pair $(i,j)$ is defined as:

$$\ell_{i,j} = -\log \frac{\exp\left(\mathrm{sim}\left(\mathrm{z}_i^{(1)},\mathrm{z}_i^{(2)}\right)/\tau\right)}{\sum_{k=1}^{2N} 1_{[k\neq i}\exp\left(\mathrm{sim}\left(\mathrm{z}_i^{(1)},\mathrm{z}_k\right)/\tau\right)}$$

Where $\tau$ is a temperature parameter that scales the logits before applying the softmax function and $1_{[k\neq i]}$ is an indicator function that is 1 if $k \neq i$ and 0 otherwise.[2] This loss is essential for training the CIPHER to learn representations that are robust to augmentations and generalize well to unseen data.

#### *L1 Loss*

The L1 loss, or mean absolute error (MAE), measures the difference between the original 3D CT volume block and its reconstructed version. For a 3D CT volume block **x** and its reconstruction **x̂**, the L1 loss is defined as:

$$\mathcal{L}_{\mathrm{L1}} = \frac{1}{n}\sum_{i=1}^{n} |x_i - \hat{x}_i|$$

Where $n$ represents the number of voxels in the 3D CT volume and $x_i$ and $\hat{x}_i$ denote the original and reconstructed voxel values, respectively. The L1 loss penalizes deviations between the original and reconstructed voxel values, encouraging the model to produce reconstructions that closely match the original 3D CT volumes. This enhances the overall quality of the learned representations, especially during the fine-tuning phase on our in-house dataset.

*B. Augmentation Methods*

Following the masked autoencoder principle[3], we applied advanced augmentation techniques on 3D CT volume blocks, aimed at enhancing the model's generalizability and robustness across different lung pathologies. Specifically, we employed three augmentation strategies: outer-cutout, inner-cutout, and local block shuffling. Outer-cutout removed only the extreme peripheral margins of the CT volume while retaining the chest wall, whereas inner-cutout challenged the model by removing parts of the internal thoracic anatomy. Local block shuffling was introduced to encourage the model to learn spatial relationships and contextual information within the lung tissue. This approach allows for robust feature extraction without reliance on extensively labeled datasets. The ViTAutoEnc, a vision transformer-based masked autoencoder, encodes these augmented views and reconstructs the original 3D blocks, ensuring robust feature learning.

**Supplemental Results**

In the MDA internal test cohort, CIPHER achieved an AUC of 0.827, 95% CI 0.663-0.990, compared with the clinical model, AUC 0.580, 95% CI 0.367-0.793; radiomics model, AUC 0.773, 95% CI 0.592-0.955; and ensemble model, AUC 0.778, 95% CI 0.605-0.951. Pairwise DeLong comparisons showed that CIPHER had a higher AUC than the clinical model, AUC difference 0.247, 95% CI −0.026 to 0.520, p = 0.0759; radiomics model, AUC difference 0.054, 95% CI −0.163 to 0.271, p = 0.6257; and ensemble model, AUC difference 0.049, 95% CI −0.092 to 0.190, p = 0.4962 (**Suppl Table S3**).

In the JHU external validation cohort, CIPHER achieved an AUC of 0.826, 95% CI 0.713-0.938, compared with the clinical model, AUC 0.588, 95% CI 0.437-0.738; radiomics model, AUC 0.663,

95% CI 0.551-0.776; and ensemble model, AUC 0.796, 95% CI 0.695-0.897. Pairwise DeLong comparisons showed that CIPHER significantly outperformed the clinical model, AUC difference 0.238, 95% CI 0.025-0.451, $p = 0.0287$, and the radiomics model, AUC difference 0.163, 95% CI 0.014-0.312, $p = 0.0318$. CIPHER and the ensemble model had comparable AUCs, AUC difference 0.030, 95% CI −0.082 to 0.142, $p = 0.5996$ (**Suppl Table S3**).

### Supplemental Tables

**Suppl Table S1:** CIPHER model architecture, training configuration, and runtime details.

| Item | Specification |
|---|---|
| Input patch/sub-volume size | 96 × 96 × 96 voxels |
| Masking ratio | Randomly sampled 0% to 30% voxel/block dropout per input; expected average approximately 15% |
| Mask/block size | Dropped 3D blocks sampled up to 25% of each spatial dimension, with minimum block size of 5% |
| Backbone architecture | VIT (3D Swin Transformer) self-supervised encoder |
| Transformer token patch size | 2 × 2 × 2 |
| Embedding/feature size | 48 |
| Window size | 7 × 7 × 7 |
| Depths | [2, 2, 2, 2] |
| Attention heads | [3, 6, 12, 24] |
| MLP ratio | 4.0 |
| Trainable parameters | 29,672,551 trainable parameters, approximately 29.7 million |
| Self-supervised pretraining duration | 1000 epochs |
| Fine-tuning duration | 300 epochs |
| Optimizer | AdamW |
| Learning rate | $6 \times 10^{-6}$ |
| Weight decay | 0.1 |
| Learning-rate schedule | Warmup cosine schedule with 500 warmup steps |
| Augmentation strategy | Random spatial crop; 90° in-plane rotation; outer-cutout; inner-cutout; and local 3D patch shuffling. |
| Mixed precision | Yes, automatic mixed precision enabled |
| Training hardware | 8 NVIDIA A100 GPUs |
| Average runtime per patient | 93.2 seconds per patient on a single NVIDIA A100 40 GB GPU |

**Suppl Table S2**: CIPHER foundation model results for resampling strategy repeated five times on the MDA held-out cohort

| Training Fold | Balanced Accuracy | Sensitivity | Specificity | F1 Score | AUC | PPV | NPV |
|---|---|---|---|---|---|---|---|
| Fold 1 | 0.840 | 0.848 | 0.833 | 0.788 | 0.814 | 0.737 | 0.909 |
| Fold 2 | 0.835 | 0.787 | 0.883 | 0.787 | 0.766 | 0.788 | 0.883 |
| Fold 3 | 0.831 | 0.878 | 0.783 | 0.773 | 0.835 | 0.690 | 0.922 |
| Fold 4 | 0.867 | 0.818 | 0.916 | 0.830 | 0.845 | 0.844 | 0.902 |
| Fold 5 | 0.840 | 0.848 | 0.833 | 0.788 | 0.829 | 0.737 | 0.909 |

**Suppl Table S3:** Pairwise DeLong comparisons of AUCs.

| Cohort | Comparison | Model 1 AUC (95% CI) | Model 2 AUC (95% CI) | AUC difference | 95% CI for AUC difference | DeLong p-value |
|---|---|---|---|---|---|---|
| MDA internal benchmark cohort | Radiomics vs Clinical | 0.773 (0.592-0.955) | 0.580 (0.367-0.793) | 0.193 | -0.029 to 0.415 | 0.0889 |
| | CIPHER vs Clinical | 0.827 (0.663-0.990) | 0.580 (0.367-0.793) | 0.247 | -0.026 to 0.520 | 0.0759 |
| | Ensemble vs Clinical | 0.778 (0.605-0.951) | 0.580 (0.367-0.793) | 0.198 | 0.017 to 0.379 | 0.0323 |
| | CIPHER vs Radiomics | 0.827 (0.663-0.990) | 0.773 (0.592-0.955) | 0.054 | -0.163 to 0.271 | 0.6257 |
| | Ensemble vs Radiomics | 0.778 (0.605-0.951) | 0.773 (0.592-0.955) | 0.005 | -0.166 to 0.176 | 0.9544 |
| | CIPHER vs Ensemble | 0.827 (0.663-0.990) | 0.778 (0.605-0.951) | 0.049 | -0.092 to 0.190 | 0.4962 |
| JHU external validation cohort | Radiomics vs Clinical | 0.663 (0.551-0.776) | 0.588 (0.437-0.738) | 0.075 | -0.106 to 0.256 | 0.4169 |
| | CIPHER vs Clinical | 0.826 (0.713-0.938) | 0.588 (0.437-0.738) | 0.238 | 0.025 to 0.451 | 0.0287 |
| | Ensemble vs Clinical | 0.796 (0.695-0.897) | 0.588 (0.437-0.738) | 0.208 | 0.059 to 0.357 | 0.0062 |
| | CIPHER vs Radiomics | 0.826 (0.713-0.938) | 0.663 (0.551-0.776) | 0.163 | 0.014 to 0.312 | 0.0318 |
| | Ensemble vs Radiomics | 0.796 (0.695-0.897) | 0.663 (0.551-0.776) | 0.133 | 0.048 to 0.218 | 0.0022 |
| | CIPHER vs Ensemble | 0.826 (0.713-0.938) | 0.796 (0.695-0.897) | 0.030 | -0.082 to 0.142 | 0.5996 |

**Abbreviations**: AUC, area under the receiver operating characteristic curve; CI, confidence interval; JHU, Johns Hopkins University; MDA, MD Anderson Cancer Center. AUC differences are calculated as Model 1 minus Model 2. AUC confidence intervals and pairwise comparisons were estimated using DeLong's method.[4]

**Suppl Table S4:** Calibration and predictive performance for clinical, radiomics, CIPHER, and ensemble models.

| Cohort | Model | ICI-P events/N (%) | AUC | Brier score | Calibration intercept | Calibration slope | Sensitivity | Specificity | PPV | NPV |
|---|---|---|---|---|---|---|---|---|---|---|
| MDA internal benchmark cohort | Clinical | 15/30 (50.0) | 0.580 | 0.248 | -0.024 | 0.568 | 0.800 | 0.400 | 0.571 | 0.667 |
| | Radiomics | 15/30 (50.0) | 0.773 | 0.200 | -0.205 | 1.713 | 0.733 | 0.800 | 0.786 | 0.750 |
| | CIPHER | 15/30 (50.0) | 0.827 | 0.187 | 0.134 | 2.015 | 0.800 | 0.867 | 0.857 | 0.813 |
| | Ensemble | 15/30 (50.0) | 0.778 | 0.203 | -0.069 | 2.913 | 0.800 | 0.800 | 0.800 | 0.800 |
| JHU external validation | Clinical | 20/116 (17.2) | 0.587 | 0.215 | -1.437 | 0.574 | 0.500 | 0.781 | 0.323 | 0.882 |
| | Radiomics | 20/116 (17.2) | 0.663 | 0.237 | -1.600 | 0.848 | 0.850 | 0.458 | 0.246 | 0.936 |
| | CIPHER | 20/116 (17.2) | 0.826 | 0.175 | -1.453 | 2.284 | 0.800 | 0.833 | 0.500 | 0.952 |
| | Ensemble | 20/116 (17.2) | 0.796 | 0.194 | -1.277 | 3.426 | 0.900 | 0.625 | 0.333 | 0.968 |

**Note:** Calibration and performance metrics were calculated using original model probabilities and the locked classification threshold.

**Suppl Table S5:** Prevalence-adjusted PPV and NPV across clinically plausible ICI-P prevalence levels. PPV and NPV were recalculated across assumed ICI-P prevalence levels using the locked classification threshold and the sensitivity and specificity reported in Table 2. These values illustrate the prevalence dependence of predictive values and should not be interpreted as recalibrated absolute risk estimates.

| Model | Assumed ICI-P prevalence | MDA internal benchmark cohort | | JHU external validation cohort | |
|---|---|---|---|---|---|
| | | Prevalence-adjusted PPV | Prevalence-adjusted NPV | Prevalence-adjusted PPV | Prevalence-adjusted NPV |
| Clinical | 5.0% | 0.066 | 0.974 | 0.107 | 0.967 |
| | 9.5% | 0.123 | 0.950 | 0.193 | 0.937 |
| | 10.0% | 0.129 | 0.947 | 0.202 | 0.934 |
| | 15.0% | 0.190 | 0.919 | 0.287 | 0.898 |
| | 17.2% | 0.217 | 0.906 | 0.322 | 0.883 |
| | 20.0% | 0.250 | 0.889 | 0.363 | 0.862 |
| Radiomics | 5.0% | 0.162 | 0.983 | 0.076 | 0.983 |
| | 9.5% | 0.278 | 0.966 | 0.141 | 0.967 |
| | 10.0% | 0.289 | 0.964 | 0.148 | 0.965 |
| | 15.0% | 0.393 | 0.944 | 0.217 | 0.945 |
| | 17.2% | 0.432 | 0.935 | 0.246 | 0.936 |
| | 20.0% | 0.478 | 0.923 | 0.282 | 0.924 |

| Model | Assumed ICI-P prevalence | MDA internal benchmark cohort | | JHU external validation cohort | |
|---|---|---|---|---|---|
| | | Prevalence-adjusted PPV | Prevalence-adjusted NPV | Prevalence-adjusted PPV | Prevalence-adjusted NPV |
| CIPHER | 5.0% | 0.240 | 0.988 | 0.201 | 0.988 |
| | 9.5% | 0.387 | 0.976 | 0.335 | 0.975 |
| | 10.0% | 0.401 | 0.975 | 0.347 | 0.974 |
| | 15.0% | 0.515 | 0.961 | 0.458 | 0.959 |
| | 17.2% | 0.555 | 0.954 | 0.499 | 0.952 |
| | 20.0% | 0.601 | 0.945 | 0.545 | 0.943 |
| Ensemble | 5.0% | 0.174 | 0.987 | 0.112 | 0.992 |
| | 9.5% | 0.296 | 0.974 | 0.201 | 0.983 |
| | 10.0% | 0.308 | 0.973 | 0.211 | 0.983 |
| | 15.0% | 0.414 | 0.958 | 0.298 | 0.973 |
| | 17.2% | 0.454 | 0.951 | 0.333 | 0.968 |
| | 20.0% | 0.500 | 0.941 | 0.375 | 0.962 |

**Abbreviations:** ICI-P, immune checkpoint inhibitor-induced pneumonitis; JHU, Johns Hopkins University; MDA, MD Anderson Cancer Center; NPV, negative predictive value; PPV, positive predictive value.

**Suppl Table S6**: Sensitivity analysis excluding patients with known baseline ILD or documented baseline pulmonary abnormalities in the MDA internal held-out cohort

| Analysis | N | ICI-P events | Non-ICI-P | Excluded N | Excluded ICI-P | Excluded non-ICI-P | AUC | Accuracy | Balanced accuracy | Sensitivity | Specificity | PPV | NPV | F1 |
|---|---|---|---|---|---|---|---|---|---|---|---|---|---|---|
| Primary internal held-out cohort | 93 | 33 | 60 | 0 | 0 | 0 | 0.845 | 0.849 | 0.849 | 0.848 | 0.85 | 0.757 | 0.911 | 0.8 |
| Excluding known baseline ILD | 91 | 31 | 60 | 2 | 2 | 0 | 0.839 | 0.846 | 0.844 | 0.839 | 0.85 | 0.743 | 0.911 | 0.788 |
| Excluding known ILD or restrictive/combined impairment | 90 | 31 | 59 | 3 | 2 | 1 | 0.839 | 0.844 | 0.843 | 0.839 | 0.847 | 0.743 | 0.909 | 0.788 |
| Excluding known ILD or documented baseline abnormality | 58 | 17 | 41 | 35 | 16 | 19 | 0.762 | 0.81 | 0.797 | 0.765 | 0.829 | 0.65 | 0.895 | 0.703 |

**Suppl Table S7**: Sensitivity analysis excluding patients with recent respiratory infection, baseline fever, or baseline respiratory symptoms in the MDA internal held-out cohort.

| Analysis | N | ICI-P events | Non-ICI-P | Excluded (N) | Excluded ICI-P | Excluded non-ICI-P | AUC | Accuracy | Balanced accuracy | Sensitivity | Specificity | PPV | NPV | F1 |
|---|---|---|---|---|---|---|---|---|---|---|---|---|---|---|
| Primary internal held-out cohort | 93 | 33 | 60 | 0 | 0 | 0 | 0.845 | 0.849 | 0.849 | 0.848 | 0.850 | 0.757 | 0.911 | 0.800 |
| Excluding recent respiratory infection | 89 | 30 | 59 | 4 | 3 | 1 | 0.851 | 0.865 | 0.866 | 0.867 | 0.864 | 0.765 | 0.927 | 0.813 |
| Excluding baseline fever | 92 | 32 | 60 | 1 | 1 | 0 | 0.841 | 0.848 | 0.847 | 0.844 | 0.850 | 0.750 | 0.911 | 0.794 |
| Excluding any baseline respiratory symptom | 58 | 18 | 40 | 35 | 15 | 20 | 0.853 | 0.879 | 0.867 | 0.833 | 0.900 | 0.789 | 0.923 | 0.811 |
| Excluding recent infection, fever, or any baseline respiratory symptom | 55 | 16 | 39 | 38 | 17 | 21 | 0.838 | 0.891 | 0.868 | 0.813 | 0.923 | 0.813 | 0.923 | 0.813 |

**Suppl Table S8**: Model performance by CT contrast status in the JHU external validation cohort

| Contrast status | Model | N | ICI-P events | AUC | 95% CI | Sensitivity | Specificity | PPV | NPV | Interaction p-value |
|---|---|---|---|---|---|---|---|---|---|---|
| Contrast-enhanced | Clinical | 98 | 19 | 0.601 | 0.452-0.742 | 0.526 | 0.658 | 0.270 | 0.852 | 0.066 |
| | Radiomics | 98 | 19 | 0.643 | 0.537-0.762 | 0.526 | 0.671 | 0.278 | 0.855 | 0.313 |
| | CIPHER | 98 | 19 | 0.826 | 0.704-0.935 | 0.789 | 0.722 | 0.405 | 0.934 | 0.456 |
| | Ensemble | 98 | 19 | 0.793 | 0.682-0.894 | 0.789 | 0.709 | 0.395 | 0.933 | 0.412 |
| Non-contrast | Clinical | 18 | 1 | 0.118 | 0.000-0.294 | 0.000 | 0.765 | 0.000 | 0.929 | 0.066 |
| | Radiomics | 18 | 1 | 0.941 | 0.812-1.000 | 1.000 | 0.647 | 0.143 | 1.000 | 0.313 |
| | CIPHER | 18 | 1 | 0.941 | 0.812-1.000 | 1.000 | 0.824 | 0.250 | 1.000 | 0.456 |
| | Ensemble | 18 | 1 | 0.941 | 0.812-1.000 | 1.000 | 0.824 | 0.250 | 1.000 | 0.412 |

**Note:** AUC, area under the receiver operating characteristic curve; CI, confidence interval; ICI-P, immune checkpoint inhibitor-associated pneumonitis; NPV, negative predictive value; PPV, positive predictive value. AUC 95% CIs were estimated using percentile bootstrap resampling. Interaction p-values were obtained from likelihood-ratio tests comparing logistic regression models with and without the model score by contrast-status interaction term. Classification metrics were calculated using the prespecified 0.5 threshold. Because the non-contrast subgroup contained only one ICI-P event, estimates for this subgroup should be interpreted cautiously.

**Suppl Table S9**: Exploratory CIPHER performance by CTCAE pneumonitis grade in the MDA internal held-out and JHU external cohorts. Detection rate was calculated among grade-labeled ICI-P cases using the locked CIPHER threshold. Exact binomial 95% confidence intervals are shown for grade-specific detection rates. Median CIPHER predicted probability and reconstruction MSE are reported with interquartile ranges. This exploratory analysis assessed whether the binary CIPHER output was associated with CTCAE pneumonitis severity and was not intended to evaluate multiclass grade prediction.

| MDA Internal Cohort | | | | | | |
|---|---|---|---|---|---|---|
| **CTCAE pneumonitis grade** | **N** | **Detected as high risk** | **Missed** | **Detection rate % (95% CI)** | **Median CIPHER probability (IQR)** | **Median CIPHER MSE (IQR)** |
| Grade 2 | 16 | 13 | 3 | 81.2 (54.4–96.0) | 0.624 (0.618–0.633) | 0.0437 (0.0213–0.0577) |
| Grade 3 | 12 | 10 | 2 | 83.3 (51.6–97.9) | 0.625 (0.613–0.631) | 0.0335 (0.0163–0.0675) |
| Grade 4 | 1 | 1 | 0 | 100.0 (2.5–100.0) | 0.646 (0.646–0.646) | 0.0320 (0.0320–0.0320) |
| Grade 5 | 4 | 4 | 0 | 100.0 (39.8–100.0) | 0.638 (0.637–0.641) | 0.0420 (0.0294–0.0581) |
| **JHU External Cohort** | | | | | | |

| | | | | | | |
|---|---|---|---|---|---|---|
| Grade 2 | 6 | 4 | 2 | 66.7 (22.3–95.7) | 0.624 (0.427–0.626) | 0.1013 (0.0226–0.1375) |
| Grade 3 | 8 | 8 | 0 | 100.0 (63.1–100.0) | 0.631 (0.619–0.636) | 0.0993 (0.0812–0.1285) |
| Grade 4 | 2 | 2 | 0 | 100.0 (15.8–100.0) | 0.642 (0.639–0.645) | 0.0769 (0.0660–0.0879) |
| Grade 5 | 4 | 2 | 2 | 50.0 (6.8–93.2) | 0.633 (0.579–0.638) | 0.0249 (0.0066–0.0580) |

**Abbreviations**: CI, confidence interval; CTCAE, Common Terminology Criteria for Adverse Events; ICI-P, immune checkpoint inhibitor-associated pneumonitis; IQR, interquartile range; MDA, MD Anderson Cancer Center; MSE, mean squared error.

### Supplemental Figures

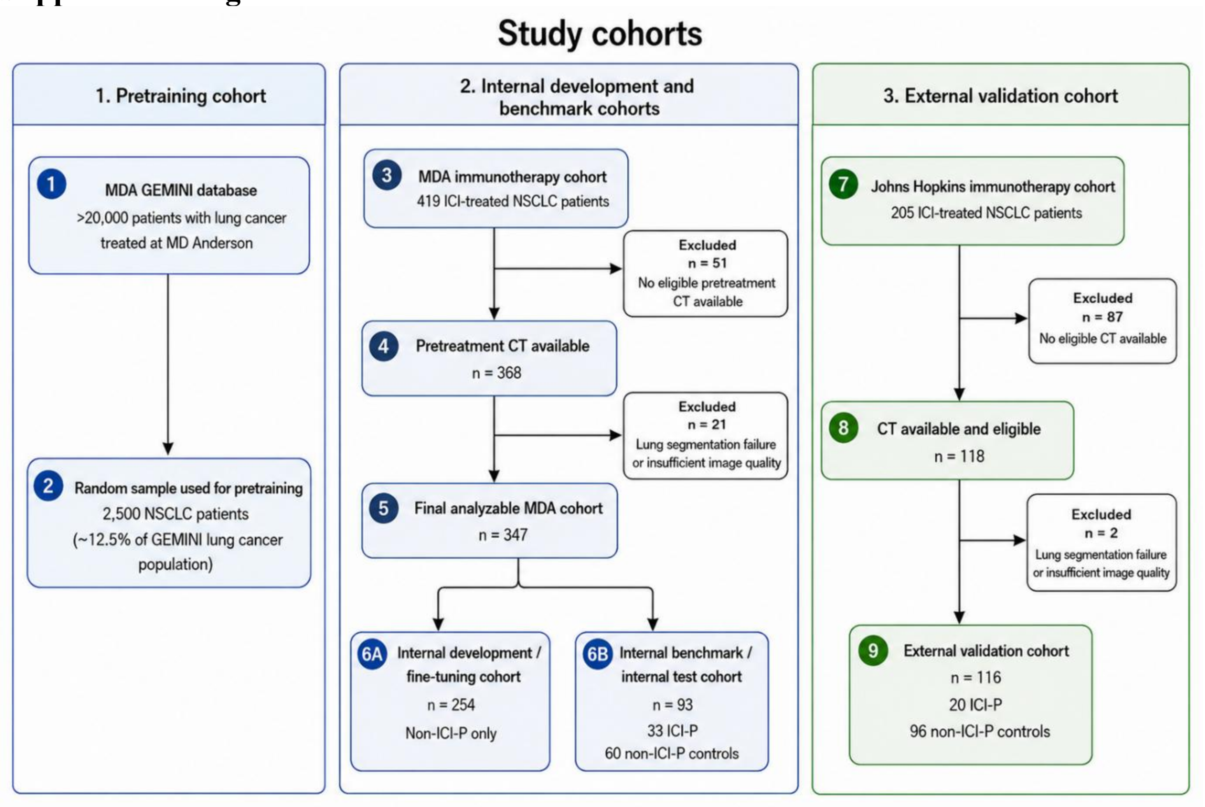


**Suppl Figure S1:** Study cohort flow diagram. The diagram summarizes the source populations, exclusions, and analytic cohorts used for CIPHER pretraining, internal development, internal testing/benchmarking, and external validation.

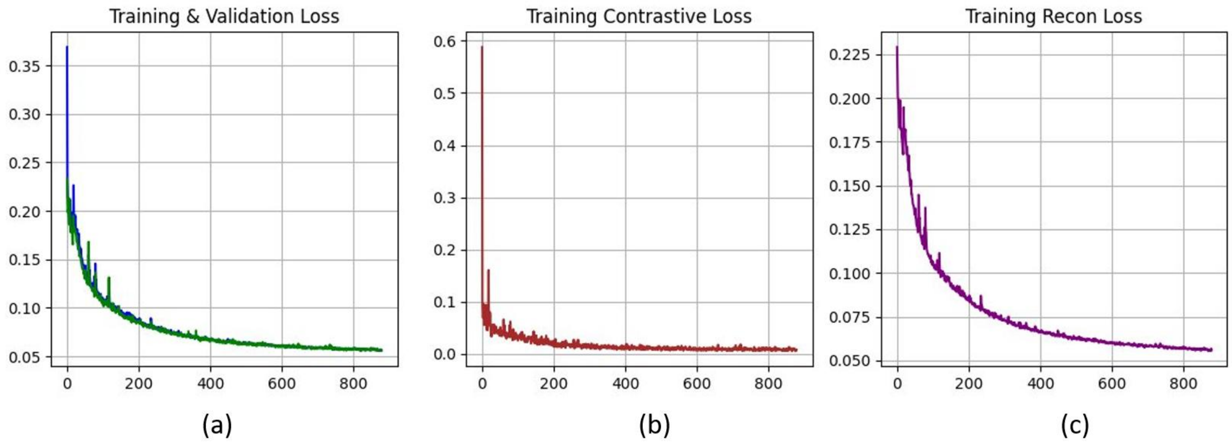


**Suppl Figure S2:** Graphical representation of training dynamics over 1000 epochs: (a) training and validation loss, (b) contrastive loss, and (c) reconstruction loss. These graphs depict the model's progressive learning and stabilization.

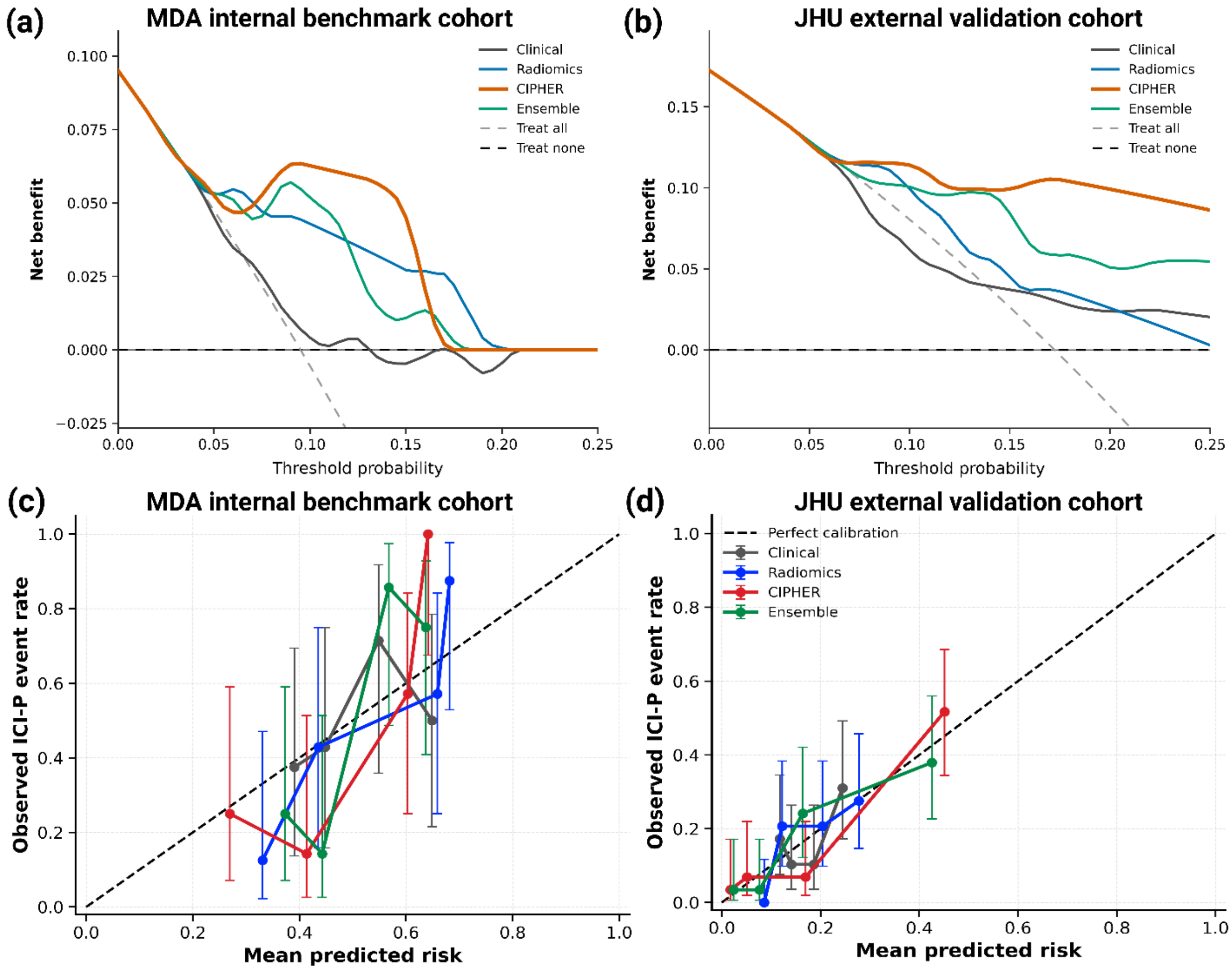


**Suppl Figure S3:** Decision-curve analysis and calibration diagnostics for clinical, radiomics, CIPHER, and ensemble models. Panels a and b show decision-curve analysis in the MDA internal benchmark cohort and Johns Hopkins external validation cohort, respectively. Net benefit is shown relative to the treat-all and treat-none reference strategies. CIPHER demonstrated the most favorable net-benefit profile among evaluated models across most clinically relevant threshold probabilities in both cohorts. Panels c and d show calibration plots for the MDA and Johns Hopkins cohorts. Error bars represent 95% Wilson confidence intervals.

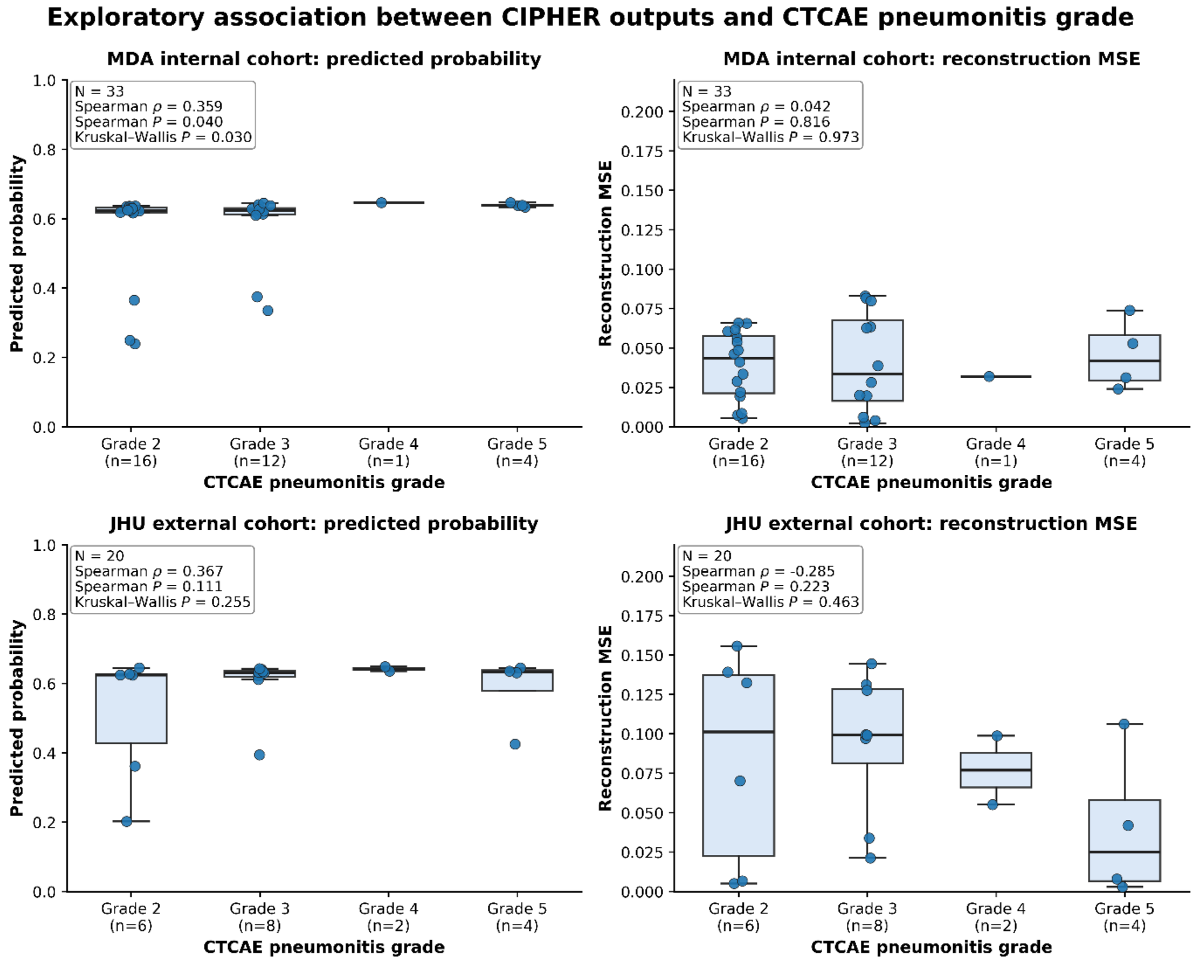


**Suppl Figure S4**: Exploratory association between CIPHER outputs and CTCAE pneumonitis grade. Boxplots show CIPHER predicted probability and reconstruction MSE across CTCAE pneumonitis grades in the MDA internal held-out cohort and Johns Hopkins external validation cohort. Individual points represent patients; boxes indicate the median and interquartile range, and whiskers extend to the most extreme values within 1.5 times the interquartile range. For grade categories containing a single patient, only the individual observation is informative because an interquartile range cannot be estimated. Annotations report sample size, Spearman correlation, Spearman p-value, and Kruskal-Wallis p-value. This exploratory analysis assessed whether the binary CIPHER output was associated with pneumonitis severity and was not intended to evaluate multiclass grade prediction.
